# Genomic landscape of virus-associated cancers

**DOI:** 10.1101/2023.02.14.23285775

**Authors:** Karen Gomez, Gianluca Schiavoni, Yoonhee Nam, Jean-Baptiste Reynier, Cole Khamnei, Michael Aitken, Giuseppe Palmieri, Antonio Cossu, Arnold Levine, Carel van Noesel, Brunangelo Falini, Laura Pasqualucci, Enrico Tiacci, Raul Rabadan

## Abstract

It has been estimated that 15%-20% of human cancers are attributable to infections, mostly by carcinogenic viruses. The incidence varies worldwide, with a majority affecting developing countries. Here, we present a comparative analysis of virus-positive and virus-negative tumors in nine cancers linked to five viruses. We find that virus-positive tumors occur more frequently in males and show geographical disparities in incidence. Genomic analysis of 1,658 tumors reveals virus-positive tumors exhibit distinct mutation signatures and driver gene mutations and possess a lower somatic mutation burden compared to virus-negative tumors of the same cancer type. For example, compared to the respective virus-negative counterparts, virus-positive cases across different cancer histologies had less often mutations of *TP53* and deletions of 9p21.3/*CDKN2*A-*CDKN1A*; Epstein-Barr virus-positive (EBV+) gastric cancer had more frequent mutations of *EIF4A1* and *ARID1A* and less marked mismatch repair deficiency signatures; and EBV-positive cHL had fewer somatic genetic lesions of JAK-STAT, NF-κB, PI3K-AKT and HLA-I genes and a less pronounced activity of the aberrant somatic hypermutation signature. In cHL, we also identify germline homozygosity in HLA class I as a potential risk factor for the development of EBV-positive Hodgkin lymphoma. Finally, an analysis of clinical trials of PD-(L)1 inhibitors in four virus-associated cancers suggested an association of viral infection with higher response rate in patients receiving such treatments, which was particularly evident in gastric cancer and head and neck squamous cell carcinoma. These results illustrate the epidemiological, genetic, prognostic, and therapeutic trends across virus-associated malignancies.

## Introduction

An estimated 15-20% of cancers are attributable to infections^1,2^, and 8-10% are caused by viruses^3,4^. To date, seven viruses are known to be associated with the development of cancers in humans (oncoviruses): human gammaherpesvirus 4 (HHV-4, also known as Epstein-Barr virus [EBV]), human herpesvirus 8 (HHV-8), human papillomavirus (HPV), human T-cell lymphotropic virus type 1 (HTLV-1), hepatitis B virus (HBV), hepatitis C virus (HCV), and Merkel cell polyomavirus (MCPyV)^5^. The first human oncovirus to be described was EBV, following the discovery of viral particles in cultured lymphoblasts from Burkitt lymphoma (BL) in 1964^6^. Since then, EBV has been linked to a wide array of both hematological and solid tumors, including Hodgkin lymphoma (HL) (20-50% of cases^7^), other B and T cell lymphomas (such as plasmablastic lymphoma [PBL], extranodal NK T-cell lymphomas [NKTCL], and primary central nervous system lymphoma [PCNSL] in immune-suppressed patients, each in 70-100% of cases^8–10^), gastric cancer (GC) (8.7% of cases^11^), and nasopharyngeal carcinoma (NPC) (100% of cases^12^), summing to approximately 1% of all cancers^3^. It has also been suggested that EBV may be associated to B-cell lymphomagenesis more widely than currently acknowledged, possibly via a “hit and run” mechanism^13^. By the 1980s, HPV, HBV, HCV, and HTLV-1 had been identified as additional oncoviruses. High-risk HPV types such as HPV16 and HPV18 contribute to the pathogenesis of cervical cancer (CC) (95% of cases^14^), head and neck squamous cell carcinoma (HNSCC) (30%^15^), and anogenital cancers (70-90%^16^), representing 5% of all cancers^16^. HBV and HCV have been associated with up to 56% and 20% of hepatocellular carcinomas (HCC)^17^ and 2% and 1% of total cancers^3^, respectively. HTLV-1, the only oncogenic retrovirus yet described, is necessary for the development of adult T-cell leukemia/lymphoma^18^. Since the discovery of HHV-8 in AIDS patients with Kaposi sarcoma (KS) in 1994^19^, HHV-8 has been implicated in the pathogenesis of Kaposi sarcoma (100% of cases), primary effusion lymphoma (100%^20^), and Castleman’s disease (20-40%^21^). In 2008, MCPyV was linked to Merkel cell carcinoma (MCC)^22^, and has since been identified as an etiological agent in 80% of MCC tumors^23^. Recently, HPV42, previously classified as a “low-risk” HPV type, has been found in a majority of digital papillary adenocarcinomas^24^. It is suspected that viruses may play a causative role in the pathogenesis of other cancer types^25,26^, and there may be more oncoviruses that have yet to be discovered.

While the mechanisms of malignant transformation caused by oncoviruses differ, there are some general patterns that are observed^27^. First, oncoviruses cause a persistent, long-term infection, and tumors develop years after the initial infection. For example, most individuals are infected with EBV by early childhood (in developing countries) or adolescence (in developed countries)^28^, but an EBV-associated cancer may not develop until old age. Hepatocellular carcinoma develops 10-30 years after infection with HBV or HCV^29^, and cervical cancer develops 25-30 years following infection with HPV^30^. Second, oncoviruses encode proteins that directly contribute to malignant transformation. In HPV infected cells, the E6 and E7 oncoproteins inhibit the tumor suppressors p53 and Rb, respectively^31^. The vGPCR protein encoded by HHV8 induces angiogenesis and promotes cell transformation^32^. EBV expresses different genes depending on the viral latency program. In Burkitt lymphoma, EBV expresses a type 1 latency program including the protein EBNA-1, which is necessary for the replication of viral DNA and may inhibit apoptosis^33^. In Hodgkin lymphoma, gastric carcinoma, and nasopharyngeal carcinoma, EBV expresses a type II latency program, including EBNA-1 as well as the proteins LMP1 and LMP2, which activate the NF-κB and PI3K/AKT pathways^33^. Third, Κ viral infection is necessary, but not sufficient for malignant transformation. Many oncoviruses are highly prevalent in the general population: 90-95% of people worldwide are infected with EBV^28^, 80% of individuals will acquire an HPV infection by age 45^34^, and MCPyV is detected in 80% of individuals in the general population by age 50^35^. However, only a small fraction of those infected with oncoviruses will develop cancer, suggesting additional genetic and/or environmental factors are required.

The factors that contribute to the malignant transformation of virus-infected cells remain incompletely understood, but are known to include a combination of environmental, immune, inherited, and somatic components. While many of these components have been described for individual cancer types, relatively little has been reported about the clinical and genetic factors that are common across virus-associated cancers. In this study, we investigate virus-associated oncogenesis through an integrative analysis of nine cancers for which a subset of cases are associated with five oncoviruses, using data from both published and newly collected data sets. We identify patterns of common phenotypic characteristics, somatic drivers, germline risk factors, and therapeutic responses among these malignancies. This study provides a comprehensive analysis of human cancers that develop in the context of viral infection and key factors related to their pathogenesis.

## Results

### Virus-associated cancer show unique epidemiological trends

Virus-associated cancers are known to follow unique epidemiological patterns compared to non-virus-associated cancers. For example, the age of incidence of HL follows a bimodal distribution which largely reflects two distinct histological subtypes with different etiologies: 1) nodular sclerosis, usually EBV-negative, in young adults, and 2) mixed cellularity, often EBV-positive, in older adults^36,37^. BL has been traditionally classified into two clinical variants, i.e. endemic BL (eBL) and sporadic BL (sBL), that present different demographic (eBL tend to be younger), geographic (most eBL are from equatorial Africa), virus status (nearly all eBL are EBV+ while a small fraction of sBL are) and somatic mutation characteristics^38^ (this should be taken into account in BL comparisons along the manuscript).

In order to illustrate other common demographic characteristics of virus-associated malignancies, we analyzed data from the Global Cancer Observatory (GLOBOCAN 2020)^39^ and published incidence rates in 82 studies of 11 cancer types linked to 7 viruses and 13 non-virus-associated cancers^40–121^ (**Table S2**). First, we compared the incidence rates of viral cancers in males versus females (M/F) reported in select published studies^40–121^ (**Figure 1A, Table S2**). We found that the M/F ratio reported was greater overall in virus-associated cancers compared to nonviral cancers (p=0.03, Mann Whitney U [MWU] test). Among studies that reported rates of male and female incidence for virus-positive and virus-negative tumors specifically, virus-positive cases tended to have a greater M/F ratio than virus-negative cases (p=2.4e-10). This trend was consistent when separately comparing virus-positive and virus-negative tumors of gastric cancer (p=1.04e-10) and Hodgkin lymphoma (p=0.015), whereas no difference in the M/F ratio and viral status was observed for BL although higher rates of male vs female incidence has been reported in both eBL and sBL, with ratios ranging from 2:1 to 4:1^122^. A lower M/F ratio was observed in MCPyV-positive MCC compared to virus-negative MCC. Interestingly, digital papillary adenocarcinoma, which has been recently associated to HPV42, is more frequent in males compared to females at a ratio of 4 to 1^123^.

**Figure 1.**
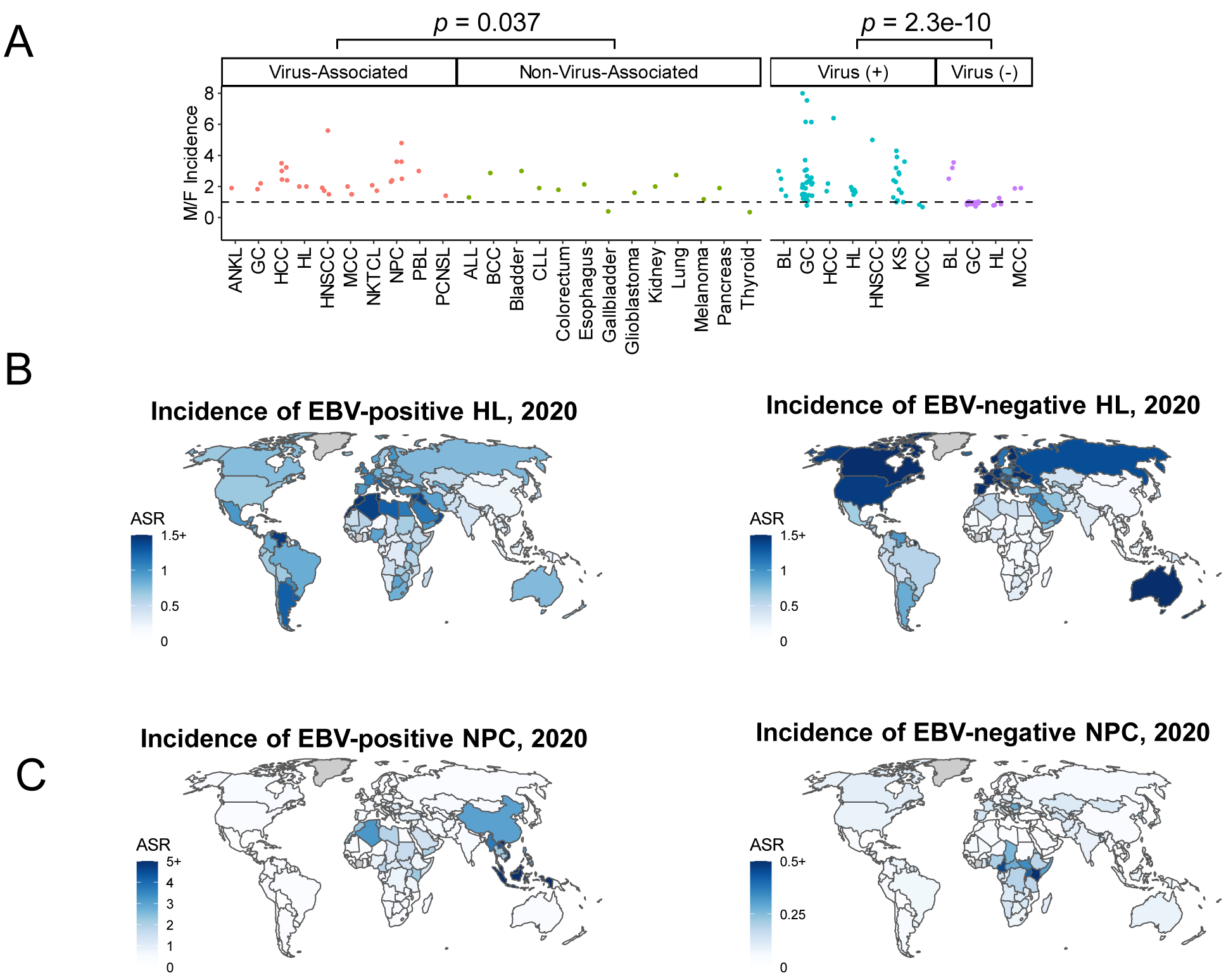
Epidemiological trends of virus-associated cancers. A) Incidence rates of virus-associated and non-virus-associated cancers (left) and virus-positive and virus-negative tumors in virus-associated cancers (right) in males compared to females (M/F) reported in selected published studies. Each point corresponds to an incidence ratio reported in a published study in Table S2. Virus-associated cancers: ANKL, aggressive NK-cell leukemia (n=1 study); GC, gastric cancer (n=2); HCC, hepatocellular carcinoma (n=6), HL, Hodgkin lymphoma (n=2); HNSCC, head and neck squamous cell carcinoma (n=4), MCC, Merkel cell carcinoma (n=3); NKTCL, Natural killer/T-cell lymphoma (n=2); NPC, nasopharyngeal carcinoma (n=6); PBL, plasmablastic lymphoma (n=1); PCNSL, primary central nervous system lymphoma (n=1). Non-virus-associated cancers (n=1 each): ALL, acute lymphoblastic leukemia; BCC, basal cell carcinoma; CLL, chronic lymphocytic leukemia. Virus-positive tumors: BL, Burkitt lymphoma (n=4); GC (n=33); HCC (n=3); HL (n=6); HNSCC (n=1); KS, Kaposi sarcoma (n=15); MCC(n=2). Virus-negative tumors: BL (n=3); GC (n=31); HL (n=6); MCC (n=2). B) Estimated incidence rates of EBV-positive HL (left) and EBV-negative HL (right) by country. C) Estimated incidence rates of EBV-positive NPC (left) and EBV-negative NPC (right) by country.

To examine how the incidence of virus-associated cancers differs by geographic location, we compared the age-standardized incidence rates (ASR) of 4 cancers in 185 countries reported in GLOBOCAN 2020. To identify countries with high ASR of virus-positive tumors for different virus-associated cancers, we estimated the number of cases attributable to viral infection by country from GLOBOCAN 2020 total incidence counts and the attributable fraction per region estimated by de Martel et al^3^. In HL, EBV-positive cases occur most frequently in North Africa, the Middle East, and South America, with the lowest incidence occurring in East Asia (**Figure 1B**). In contrast, most cases of EBV-positive NPC occur in China and southeast Asia (**Figure 1C**). Similarly, Kaposi sarcoma and cervical cancer (nearly all of which are virus-positive) show disparities in incidence by geographic location (**Figure S1**). These results illustrate that the locations of global hot spots of virus-positive tumor incidence vary by virus and even among cancers associated with the same virus. These disparities reflect differences in risk factors for virus-positive tumors among human populations, both genetic (e.g. inherited susceptibility polymorphisms^124,125^) and environmental (e.g. oncovirus prevalence^126^, and lifestyle factors such as smoking or diet, which affect overall cancer risk^127^).

### Virus-positive tumors have fewer somatic mutations than virus-negative tumors

In order to quantify the somatic mutation burden of virus-associated cancers, we aggregated somatic mutation data from 1,658 tumors in published studies of 9 cancers subjected to whole exome sequencing (classical HL^128,129^ [cHL] [n=56], PBL^130^ [n=15], GC^131^ [n=440], HCC^132^ [n=196], CC^133^ [n=178], and HNSCC^134^ [n=487]), targeted DNA sequencing (PBL^130,135^ [n=36], PCNSL^136^ [n=58], MCC^137^ [n=71], and BL^38^ [n=29]), and/or whole genome sequencing (BL^38^ [n=91] and newly sequenced cHL [n=32] [see Methods]) (**Table S1**). In general, virus-negative tumors had a higher count of nonsynonymous mutations compared to virus-positive tumors (**Figure 2A and Table S3**). The count of nonsynonymous mutations was significantly lower in virus-positive compared to virus-negative cHL (described in the next section), PCNSL (median 1 and 6, p=1.2e-7), and HNSCC (median 94.5 and 168, p=1.6e-5), and trended towards significance in PBL (median 1 and 4, p=0.080), GC (median 131.5 and 169, p=0.088), and to a lesser extent in CC (median 105 and 399, p=0.20), and MCC (median 10 and 28, p=0.17) (**Figure 2B**), whereas HCC trended in the opposite direction (median 129 in virus-positive vs 118 in virus-negative cases, p=0.12). In BL, the mutation load was significantly higher in virus-positive tumors (median 53 and 42, p=0.00018), but the count of nonsynonymous mutations in genes previously described as BL drivers^38^ trended towards lower in the virus-negative cases (median 8 and 6, p=0.074) (**Figure S2**), consistent with that report^38^. As previously mentioned, almost all EBV+ cases are eBL with unique demographic characteristics that may impact the interpretation of the results. For instance, it is known that pediatric tumors harbor less mutations than adults, which could explain the lower mutational burden of EBV+ cases. Furthermore, when restricting the analysis to driver genes, the higher mutation count in virus-negative versus virus-positive cases became statistically significant in MCC (median 5 and 1, p=0.029) (**Figure S2**;), while virus-positive GC and HCC had more driver gene mutations than their virus-negative counterparts (GC: median 3 vs 2, p=0.029; HCC: median 2 vs 1, p=0.0017). Overall, the total mutation count and/or driver mutation count was lower in virus-positive compared to virus-negative tumors in most cancers studied (**Figure 2C**).

**Figure 2:**
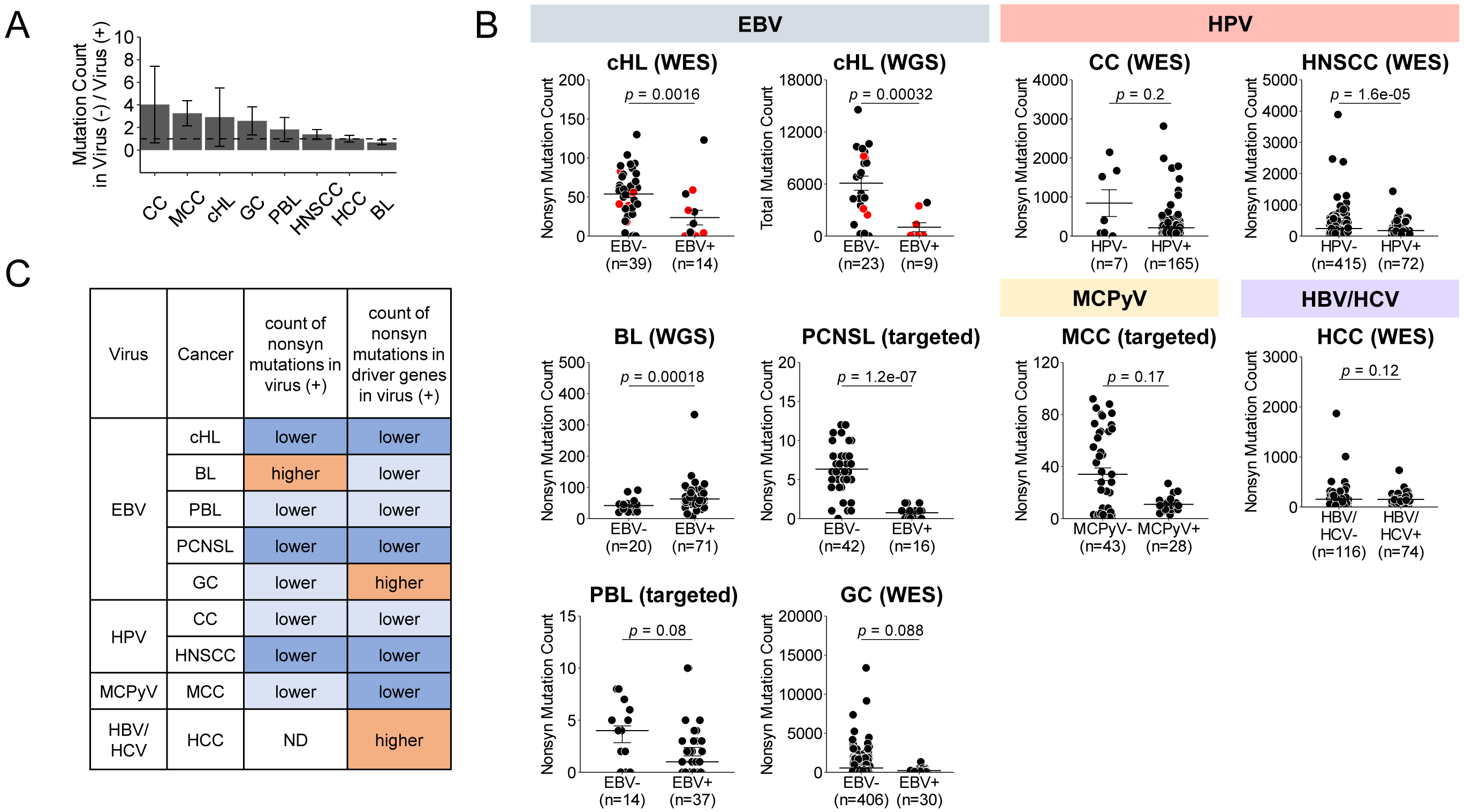
Mutation burden of virus-positive and virus-negative tumors in 9 cancers. A) Ratio of average number of somatic nonsynonymous mutations in virus-negative tumors compared to virus-positive tumors. MCC (n=71), HNSCC (n=487), CC (n=172), cHL (n=54), GC (n=436), BL (n=91; 68 EBV-positive eBL, 6 EBV-negative eBL, 3 EBV-positive sBL, 14 EBV-negative sBL), PBL (n=51), HCC (n=190), and PCNSL (n=58). B) Counts of somatic nonsynonymous mutations in virus-positive and virus-negative tumors in the same cancers. C) Summary of trends in mutation load in virus-positive versus virus-negative tumors by cancer type. Results highlighted in orange and dark blue reach significance past a threshold p <0.05, while results highlighted in light blue indicate a trend that does not reach significance p<0.05 (MWU test).

### Somatic mutations genome-wide, and alterations in JAK/STAT, NF-κB, PI3K-AKT, and HLA-I genes, are more frequent in EBV-cHL than EBV+ cHL

Hodgkin lymphoma is one of the most common malignancies in adolescents and young adults, comprising approximately 12% of all cancers in individuals aged 15-29^138^. The incidence of the EBV-positive cHL varies according to geography, and more than 50% of cHLs in developing countries are EBV-positive^7^. The genetics of EBV+ cHL has not been well studied due to the technical challenges related to the rarity of tumor cells, which usually comprise just 1-5% of cells in the tumor tissue^139^. To address this challenge, we had laser-microdissected Hodgkin Reed-Sternberg cells (n=1200-1800 per case) along with a similar number of adjacent non-neoplastic cells from frozen lymph node sections and subjected the samples to whole genome amplification (WGA) in duplicate, as previously described^128^ (see Methods). We sequenced 38 samples by WES and combined them with 18 additional cHLs from another published cohort^129^ for a total of 56 cHL, 15 of which were EBV-positive, the largest cohort of EBV-positive cHL sequenced by WES yet described. We found that the number of nonsynonymous somatic mutations in EBV-positive cHL was much lower than in EBV-negative cHL (median 4.5 compared to 57, respectively, p=0.0013, MWU test, **Figure 2B**), consistent with what we observed in other virus-positive tumors and with previous results on smaller number of cHL cases^128,129^. Importantly, because EBV-positivity is enriched in the mixed-cellularity (MC) histological subtype of cHL^36,37^, we excluded that the observed difference in somatic mutation load was associated to histology (rather than viral status) by documenting, within EBV-negative cHL, a similar somatic mutation load in the MC subtype (n=6 cases) and non-MC subtypes (n=33 cases; median of 48 and 60 nonsynonymous mutations, p=0.56).

To mitigate the potential selective pressure imposed on coding sequences we subjected to WGS 32 cHL cases (n=9 EBV-positive and 23 EBV-negative cases, of which 31 which were also included in the WES cohort). We found a considerably lower count of total clonal somatic mutations in EBV-positive than in EBV-negative cases (median of 112, range 30-3,862, and 6,826, range 9-14,564, respectively; p= 3.2e-4, MWU test, **Figure 2B**) despite similar coverage depth between these two disease groups (both median of 44X in the tumor samples [p=0.18], and 43X vs 44X in the normal samples, respectively [p=0.23]). The same pattern was found when restricting the analysis to mutations in coding regions (median of 1, range 0-19, and 50, range 0-100, respectively; p=1.2e-3, **Figure S3A**) or to nonsilent mutations (median of 1, range 0-16, and 33, range 0-82, respectively; p=9.6e-4, **Figure S3B**), consistent with the results obtained from WES data. In contrast, the fraction of short insertions/deletions among all mutation types was greater in the EBV-positive versus EBV-negative cases, suggesting potentially different underlying mechanisms (median of 22% vs 13%, respectively; p=0.024, **Figure S3C**).

Next, we investigated whether EBV-positive and EBV-negative cases cHL are preferentially associated with distinct genetic alterations, by examining the mutation frequencies of key Hodgkin lymphoma driver genes in WES data from 56 cHL cases (n=15 EBV-positive and 41 EBV-negative) (**Figure 3A**). Consistent with previous studies^140,141^, we found the JAK-STAT pathway members *STAT6* and *SOCS1* as the most frequently mutated genes, being affected in 24/56 cases (43%), the majority being EBV-negative (22/24, 92%). In particular, *STAT6* missense mutations of the DNA binding domain were observed in 14/41 EBV-negative cases (34%) but in only 1/15 EBV-positive cases (7%; p=0.047, MWU test) (**Figure 3B,C**). Furthermore, gains or amplifications of 9p24.1/*JAK2* were significantly enriched in EBV-negative cases (31/41, 76%, versus 6/15 EBV-positive cases, 40%; p=0.024). There were also mutations in the JAK-STAT gene *CSF2RB* in 4 cases, 3/4 of which were EBV-negative. Overall, at least 1 of these 4 JAK-STAT pathway genes was targeted by genetic lesions in 35/41 EBV-negative tumors (85%) but only 7/15 EBV-negative ones (47%; p=0.0057). Another frequently targeted pathway was NF-κB, with recurrent mutations and/or deletions of *TNFAIP3* at 6q23.3, mutations of *NFKBIE* and/or gain/amplification of *REL* at 2p16.1 being observed in 42/56 cases overall (75%) and more often in EBV-negative cases (35/41, 85%) than EBV-positive cases (7/15, 47%; p=0.0057). A third pathway more frequently mutated in EBV-negative tumors was PI3K-AKT (14/41, 34% EBV-cases vs 1/15, 7% EBV+ cases; p=0.047), including the pathway inhibitor genes *GNA13* and *ITPKB*. In particular, at least one nonsilent mutation in *GNA13* (missense or truncating events distributed throughout the coding sequence) or out-of-frame tandem duplication event was detected in 10/41 EBV-negative cases (24%), while none of the 15 EBV-positive cases had *GNA13* mutations (p=0.048). The lack of *GNA13* nonsilent mutations in EBV-positive cHL was further confirmed by targeted Sanger sequencing of an additional 18 EBV-positive cases (i.e., 33 in total) (p=0.0035) (**Figure 3D,E**). Finally, mutations in MHC-I genes (*B2M* and HLA class I A, B or C) were more frequent in EBV-negative (23/41, 56%) compared to EBV-positive (3/15, 20%) cases (p=0.032).

Copy number profiling of the 56 cHL tumors with GISTIC2.0 revealed 6 peaks of gain or amplification and 18 peaks of deletion (**Figure 3F,G and Table S5**). The burden of copy number gains was also greater in the EBV-negative cases compared to EBV-positive (median 4 and 3, p=0.0027, **Figure S4**).

**Figure 3:**
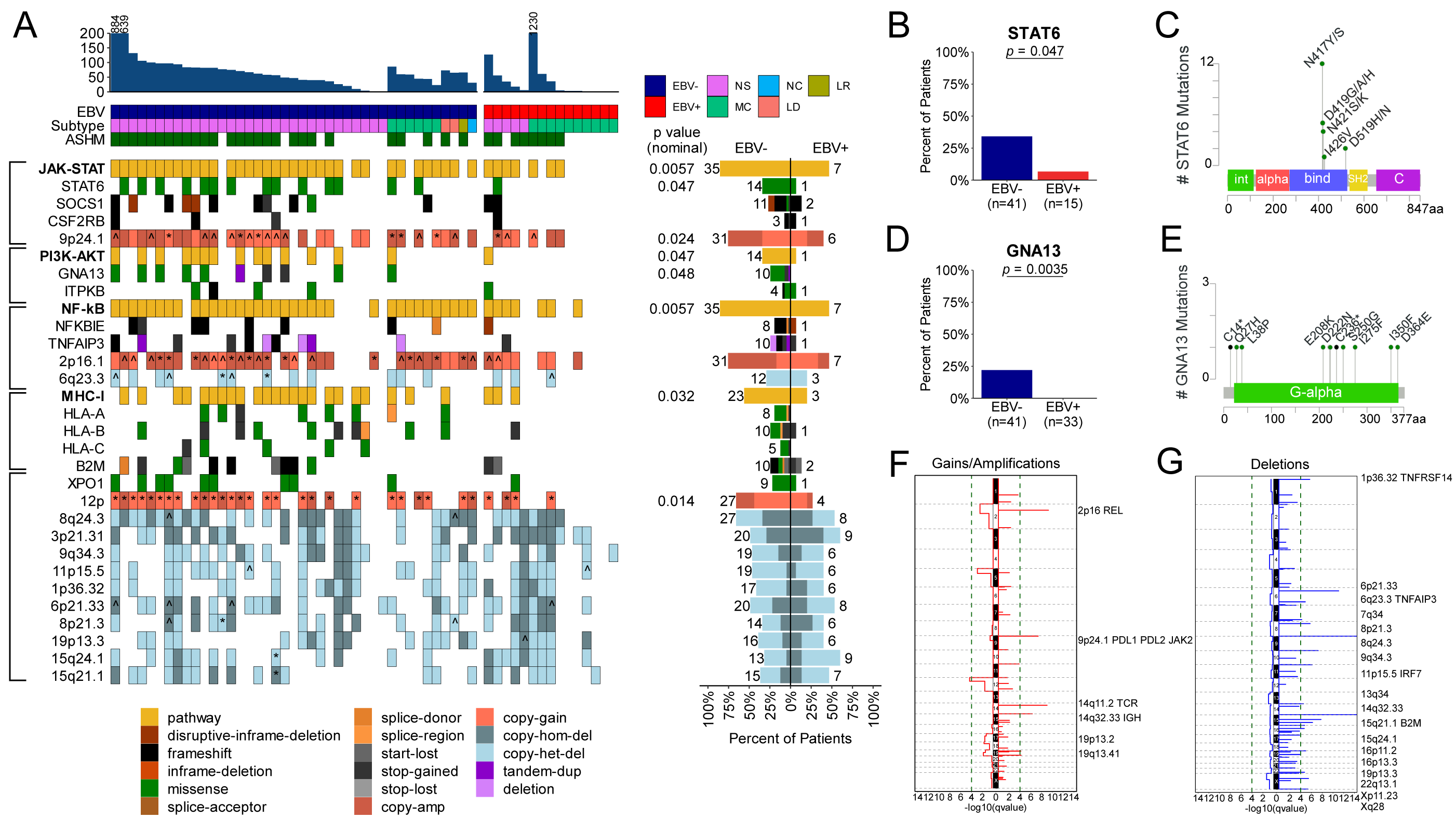
Genetic lesions in EBV-positive and EBV-negative cHL sequenced by WES. A) Genomic landscape of cHL (n=56). Clonal mutations in genes mutated in at least four patients and known to be implicated in cHL pathogenesis, as well as significant copy number peaks from GISTIC, are shown. Mutations in HLA-I genes were obtained from PolySolver, while other mutations were obtained from SAVI +/-Sanger sequencing. ^chromosomal copy number alteration; *arm-level copy number alteration. ASHM: at least one mutation in an aberrant somatic hypermutation target region. JAK-STAT: at least one mutation in STAT6, SOCS1, CSF2RB, and/or CNA in 9p24.1. PI3K-MAPK: at least one mutation in GNA13, ITPKB. NF-κB: at least one mutation in NFKBIE, TNFAIP3 and/or CNA in 6q23.3, 2p16.1. p value: Fisher’s exact test B) Counts of patients with STAT6 nonsilent mutations in EBV-(n=41) and EBV+ (n=15) cHL. P value: Fisher’s exact test. C) Nonsilent mutations in STAT6 observed in 56 cHL patients. D) Counts of patients with GNA13 nonsilent mutations in EBV-(n=41) and EBV+ (n=33) cHL. P value: Fisher’s exact test. E) Nonsilent mutations in GNA13 observed in 56 cHL patients. F) Recurrent copy number gains in cHL, identified by GISTIC (n=56). G) Recurrent copy number losses in cHL, identified by GISTIC (n=56).

### Virus-associated cancers display unique mutation signatures

To detect and quantify the relative contribution of COSMIC mutation signatures^142^ within the nine virus-associated cancers, we next applied a supervised non-negative matrix factorization approach informed by *de novo* signature calling. Virus-positive tumors exhibited different activities of mutation signatures compared to virus-negative tumors of the same cancer type (**Figure 4, Figure S5, and Tables S7, S8**).

**Figure 4:**
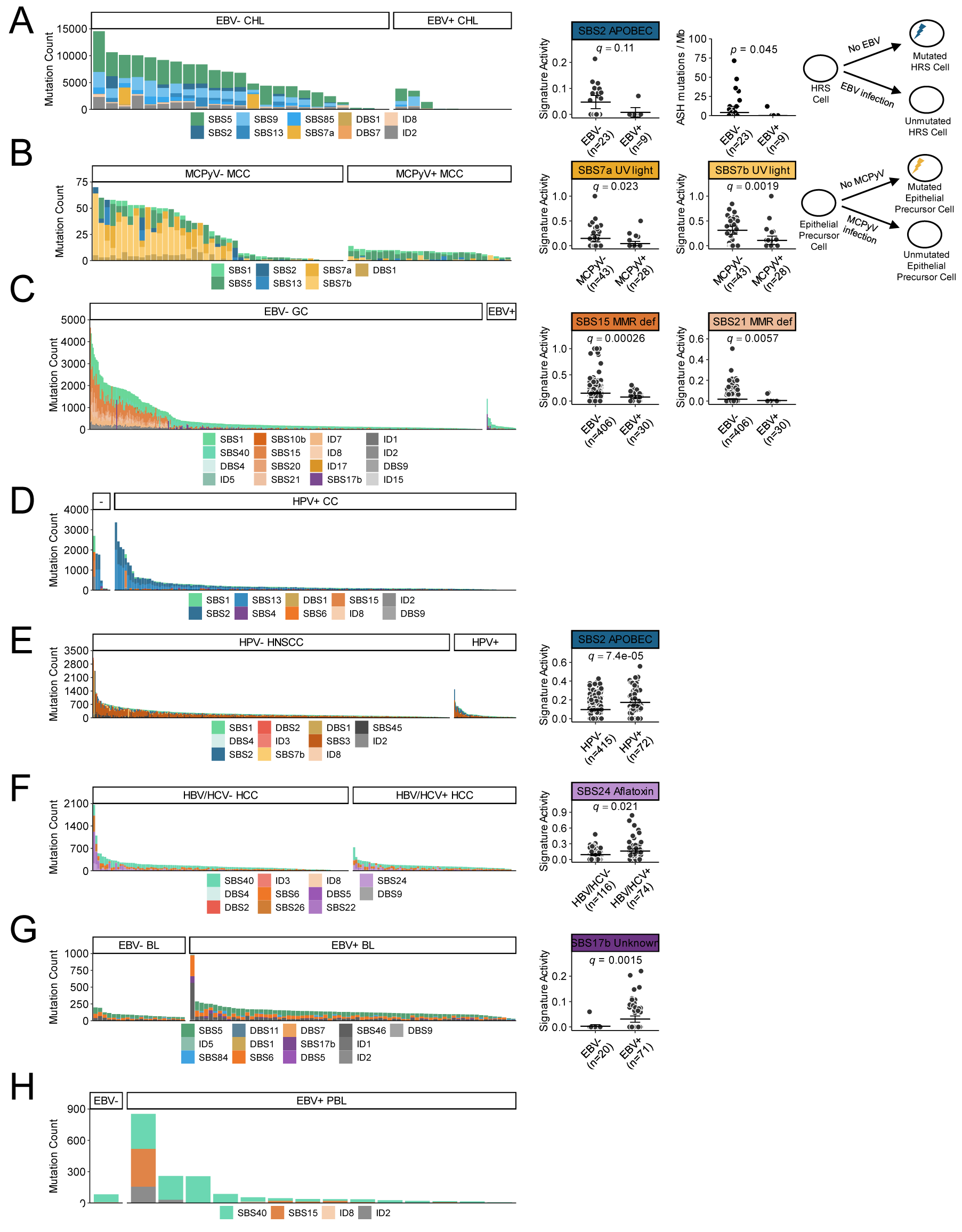
Mutations signatures in eight virus-associated cancers: A) cHL (n=32), B) MCC (n=71), C) GC (n=436), D) CC (n=172), E) HHC (n=190), F) HNSCC (n=487), G) BL (n=91; 68 EBV-positive eBL, 6 EBV-negative eBL, 3 EBV-positive sBL, 14 EBV-negative sBL), H) PBL (n=23). Activities of signatures in virus-positive compared to virus-negative cases are shown for signatures with a significant difference in activity (q<0.05) and/or biologically relevant trend (right). Schematic representation of the effect of the absence of processes behind key mutation signatures in virus-associated cases is shown in A) cHL (EBV-positive) and B) MCC (MCPyV-positive).

In cHL, the detected mutation signatures (n=10) included, among others, SBS5/age, SBS9/somatic hypermutation (SHM), SBS85/activation-induced cytidine deaminase (AID) and SBS2/13/ apolipoprotein B mRNA editing enzyme, catalytic polypeptide (APOBEC) (**Fig. 4A, left**). The absolute count of mutations ascribed to each signature was significantly lower in EBV-positive versus EBV-negative cases (**Figure S5A**). For SBS2/APOBEC, even the relative signature activity (i.e., the proportion of signature mutations among all mutations observed in each case) was lower in EBV-positive compared to EBV-negative tumors (mean proportion 0.0079 and 0.047, respectively, q=0.11) (**Figure 4A, right**). APOBEC enzymes belong to a family of cytidine deaminases that includes AID, and a previous study focused on a few selected genomic regions showed that the AID-mediated process of SHM functions aberrantly in cHL^143^. Our unbiased analysis revealed that the inferred SHM signatures SBS9 and SBS85 had higher absolute activity in EBV-negative versus EBV-positive cases genome-wide (**Figure S5A**). Additionally, we found that mutations occurring in regions within 2 kb of the transcription start site of 126 genes known to be targeted by aberrant SHM (ASHM) in diffuse large B-cell lymphomas (DLBCL)^144^ were enriched in EBV-negative compared to EBV-positive cHL (median 4 and 0 mutations per mB, p=0.045, MWU test; **Fig. 4A, right**). There were also a greater number of uniquely mutated ASHM-target genes and a greater number of mean mutations per ASHM gene in the EBV-negative than the EBV-positive patients (**Figure S6**). Collectively, these data suggest in EBV-negative cHL a more pronounced selective pressure for somatic mutations induced by APOBEC and AID activities that in EBV-positive cHL might be substituted by oncogenic activities of viral proteins. Finally, regarding the clock-like signature SBS5/age, its absolute activity did not correlate with age overall, nor separately in EBV-negative and EBV-positive cases (R2 < 0.2), in agreement with a previous report on a smaller number of cases^129^.

UV light is known to be the major etiological agent of MCC tumors in the absence of viral infection. Accordingly, the absolute count of mutations attributed to SBS7a/7b/UV light was lower in MCPyV-positive compared to MCPyV-negative (**Figure S5B**). MCPyV-positive MCC cases also displayed a lower proportion of mutations associated with each UV light signature (SBS7a/b) compared to -negative cases (mean 0.044 and 0.13, q=0.023 and mean 0.11 and 0.28, q=0.0019, respectively) (**Figure 4B**).

In GC, among mutation signatures differentially active by virus status (**Figure 4C** and **Figure S5C**), of potential interest is the greater relative and/or absolute activity of SBS20, SBS15, and SBS21 mismatch repair (MMR) deficiency signatures in EBV-negative versus EBV-positive cases. For example, mean relative activity of SBS15 is greater in EBV-negative than EBV-positive cases (mean 0.15 and 0.076, respectively; q=0.0026). These findings suggest a greater role for microsatellite instability (MSI) in the pathogenesis of EBV-negative GC. MSI as assessed by standard methods is a defining characteristic of a GC subtype that is exclusively EBV-negative and comprised 73/406 (18%) of EBV-negative patients in the TCGA cohort. Accordingly, the relative activities of SBS20, SBS15, and SBS21 were greater in the conventionally defined MSI subtype compared to the other EBV-negative cases (mean 0.11 and 0.010, p<2.2e-16; mean 0.30 and 0.12, p<2.2e-16; and mean 0.071 and 0.0071, p=4.86e-9, respectively). The absolute and relative activity of SBS15 was also greater in non-MSI EBV-negative compared to EBV-positive tumors (**Table S9**), suggesting that MSI may be more widespread in EBV-negative GC than currently appreciated with standard methods.

There was no significant difference in the absolute counts or proportions of signatures in HPV-positive versus -negative CC, likely due to the limited number (n=7) of HPV-negative cases reported in TCGA (**Figure 4D, Figure S5D**). However, in HNSCC, HPV-negative cases had a greater number of ID3 and DBS2 mutations related to smoking (**Figure S5E**), a known risk factor for HNSCC that may be less relevant for HPV-driven carcinogenesis^145^. In contrast, HPV-positive HNSCC had higher absolute and relative activity of SBS2/APOBEC (**Figure S5E and Figure 4E**), a finding consistent with the hypothesis that HPV oncoproteins may increase APOBEC3A and APOBEC3B expression and mutagenic activity^145,146^.

In HCC, there was no difference in absolute count of mutations attributed to mutation signatures (**Figure S5F),** consistent with the similar mutation burden in virus-positive and - negative HCC overall. However, HCC tumors positive for HBV and/or HCV had a greater proportion of mutations due to SBS24/aflatoxin, an environmental carcinogen known to predispose to HBV/HCV-mediated cirrhosis^147,148^ (mean 0.16 and 0.085 q=0.021) (**Figure 4F**).

In BL exomes (assessed using the reported exonic mutations from the original study^149^), we found a greater absolute and relative activity of SBS17b (a signature of unknown etiology) in EBV-positive compared to EBV-negative cases (**Figures 4G and S5G**), consistent with the original analysis of genome-wide mutation signatures in the same cohort of cases^38^. In partial contrast with that analysis, we did not detect the SBS9/polη signature associated to non-canonical AID activity that was previously found in the BL genomes^38^; and we identified in EBV-positive versus negative cases a higher absolute activity of a MMR-deficiency related signature (SBS6) distinct from that (SBS15) observed in the BL genomes^38^, as well as a higher (rather than similar^38^) load of mutations related to the clock-like signature SBS5 (whose absolute activity was not correlated with age, in the cohort overall or in EBV-positive or EBV-negative cases individually [R2<0.1]).

Overall, these results illustrate that the signatures of somatic mutation processes vary depending on infection status for each cancer, highlighting differing selective pressures on the cancer genomes in the presence or absence of viral oncoproteins.

### Virus-positive tumors harbor frequent mutations in RNA helicases *DDX3X* and *EIF4A1*

In order to identify genomic loci that are preferentially mutated in virus-positive tumors, we compared the rate of nonsynonymous mutations in the pooled cohort of 537 virus-positive tumors and 1,121 virus-negative tumors from 9 cancer types. We found that four genes had an elevated odds of mutation in virus-positive tumors compared to virus-negative tumors: *EIF4A1* (OR 69.43, 95% CI 4.15-1160.26, q=2.37e-5, MWU test, BH corrected), *DDX3X* (OR 7.07, 95% CI 4.06-12.31, q=2.25e-11), *ARID1A* (OR 2.49, 95% CI 1.72-3.58, q=7.1e-4), and *MYC* (OR 3.92, 95% CI 2.57-6.01, q=8.67e-8), although the latter was driven by GC exclusively (OR 6.13, 95% CI 0.54-70.01, q=0.38). (**Figure 5A, Table S11**). When looking at individual cancer types, *EIF4A1* had a significant OR of mutation in EBV-positive GC compared to EBV-negative GC (OR 63.09, 95% CI 2.94-1352.57, q=4.21e-6) and showed a similar trend in BL (OR 7.92, 95% CI 0.45-140.54, q=0.12) (**Figure S7, Table S11**). *DDX3X*, an RNA helicase in the same family as *EIF4A1*, had a nominally elevated OR of mutation in EBV-positive cHL (OR 8.78, 95% CI 0.34-228.59, q=0.34), HNSCC (OR 3.73, 95% CI 0.61-22.85, q=0.34), BL (OR 1.86, 95% CI 0.76-4.53, q=0.34), and GC (OR 1.64, 95% CI 0.083-32.72, q=0.74), though no cancer reached significance individually. To increase the statistical power of the association we combined data from six different BL genomic studies^38,150–154^ including eBL and sBL (n = 145 and 174, respectively) as well as EBV status (143 EBV-positive and 116 EBV-negative). We found that *DDX3X* mutation was strongly (though not exclusively) associated with EBV-positive status (OR 3.67 95% CI 2.14-6.28, p<0.001) and endemic subtype (OR 4.12 95% CI 2.55-6.67, p<0.001) (**Table S12**). *ARID1A* was significantly mutated in EBV-positive GC (OR 15.11, 95% CI 6.16-37.05, q=3.30e-12) but also trended towards an elevated OR in virus-positive HCC (OR 2.86, 95% CI 0.99-8.28, q=0.087), BL (OR 1.70, 95% CI 0.67-4.30, q=0.35), and cHL (OR 1.42, 95% CI 0.12-17.03, q=0.81). Conversely, *TP53* had an elevated odds of mutation in virus-negative tumors compared to virus-positive tumors (OR 8.58, 95% CI 6.40-11.50, q=5.8e-51) (**Figure 5A**), which was significant for most cancer types individually (**Figure S7)**. Analysis of recurrent copy number aberrations also revealed recurrent loss of 9p21.3 (*CDKN2A*, *CDKN1A*) in virus-negative tumors (**Figure S8**).

**Figure 5:**
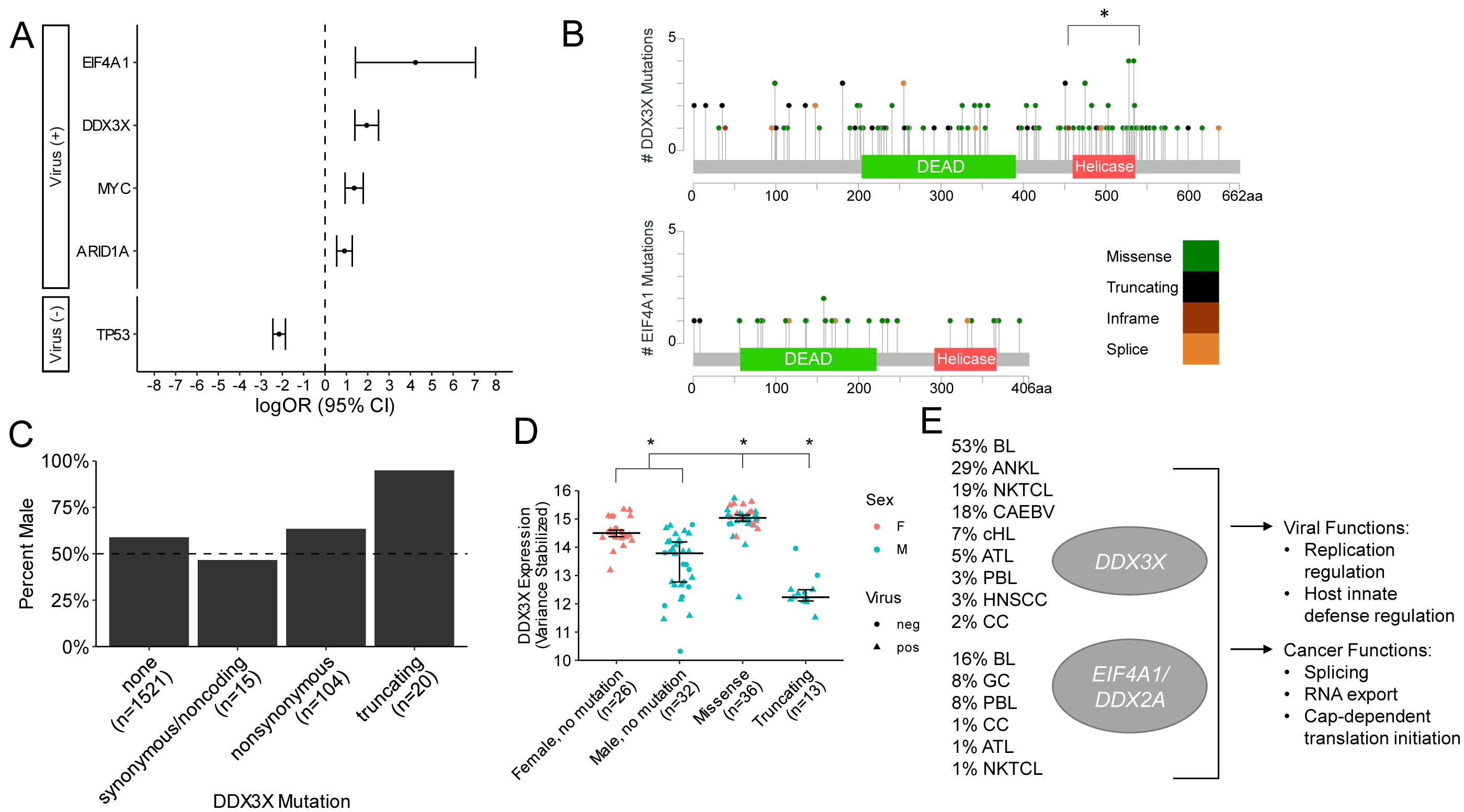
Somatic mutations in *EIF4A1* and *DDX3X* are recurrent genetic lesions associated with virus-positive status. A) Combined odds ratio of mutation in genes associated with virus-positive (top) and virus-negative (bottom) status (q<0.05) from pooled data of 1,658 tumors from 9 virus-associated cancers. *q < 0.05, Fisher’s exact test, BH corrected. B) Mutations in *DDX3X* and *EIF4A1* in 1,974 tumors. * p < 0.05, binomial test. C) Fraction of patients that are male by *DDX3X* mutation status. D) *DDX3X* expression by *DDX3X* mutation status and sex in Burkitt lymphoma (n=117). * p < 0.05, MWU test. E) Frequencies of mutation of *DDX3X* and *EIF4A1* in virus-positive tumors overall and summary of key biological functions.

*EIF4A1* and *DDX3X* are RNA helicases of the DEAD (Asp-Glu-Ala-Asp) box protein family which are known to play a role in splicing, RNA export, and cap-dependent translation initiation^155^. In order to further explore the role of these genes in virus-associated cancers, we expanded the analysis of *EIF4A1* and *DDX3X* mutations to include 316 tumors from cancers that are virus-associated in almost 100% of cases: KS (10 newly sequenced cases), aggressive NK cell lymphoma (ANKL)^156^ (n=14), ATL^157^ (n=81), NKTCL^158^ (n=100), and NPC^159^ (n=111) (**Table S1**), for a total of 1,974 cases. In all, we identified 135 DDX3X nonsynonymous mutations (100 in virus-positive; 35 in virus-negative cases) and 27 *EIF4A1* nonsynonymous mutations (22 in virus-positive; 5 in virus-negative cases).

Among virus-positive tumor samples, somatic mutations in *DDX3X* were detected in 53% (50/93) BL, 29% (4/14) ANKL, 19% (19/100) NKTCL, 7% (1/14) cHL, 5% (4/81) ATL, 3% (1/30) PBL, 3% (2/60) HNSCC, and 2% (3/144) of CC (**Table S13**). Mutations in *DDX3X* tended to occur in the helicase domain residues more frequently than expected by chance (p=4.0e-4, binomial test) suggesting selective pressure for a functional role (**Figure 5B**). This was similar for virus-positive cases only (p=0.0020) as well as virus-negative only (p=0.15). Furthermore, the proportion of mutated residues in the helicase domain was significantly greater than that of the DEAD domain (p=0.0051, Fisher’s exact test) and the region of the protein preceding the DEAD domain from amino acids 1-203 (p=0.0002, Fisher’s exact test). The *DDX3X* gene is located on the X chromosome and was previously reported to escape X inactivation in females^160^. Both truncating events and at least some missense mutations in *DDX3X* have been previously described as causing functional loss of protein activity^154^. Notably, while only 60% of patients were male (910/1529 with available data), *DDX3X* mutations that were truncating occurred almost exclusively in males (19/20, 95%) (**Figure 5C**), consistent with a previous study^161^. This was similarly evident in virus-positive (10/10, 100%) and virus-negative (9/10, 90%) cases separately. As 50% of patients with nonsynonymous *DDX3X* mutations were from the BL cohort (61/123 cases), we focused on this disease to evaluate the relationship between mutation status and *DDX3X* expression, using previously published RNA-sequencing data^38^. We found that male patients lacking somatic *DDX3X* mutations had a significantly lower expression of *DDX3X* compared to DDX3X unmutated female patients (median 13.79 and 14.50, p=2.4e-6, MWU test), consistent with escape from X inactivation^160^. Cases with missense mutations in *DDX3X* had an elevated expression of *DDX3X* irrespective of sex compared to cases with no mutations in *DDX3X* (median 15.04 and 14.24, p=1.2e-9), potentially suggesting that overexpression of missense mutants may favor their ability to decrease DDX3X function, while cases with truncating mutations (9 EBV+; 4 EBV-) had a lower expression (median 12.23 and 14.24, p=1.79e-5) (**Figure 5D**), consistent with them being loss-of-function events. Similarly, in the TCGA study of HNSCC, expression of DDX3X was lower in unmutated male cases compared to unmutated female cases (median 12.58 and 12.99, p<2.2e-16), and was also lower in cases (3 HPV+; 2 HPV-) with truncating mutations compared to unmutated cases (median 10.37 and 12.71, p=1.5e-4) (**Figure S9**). Together, these results suggest that mutations in *DDX3X* and *EIF4A1* may play a role in virus-positive tumors in various types of cancer (**Figure 5E**).

### HLA-I homozygosity is a germline risk factor for EBV-positive Hodgkin lymphoma

The major histocompatibility complex class I (MHC-I) plays an essential role in the adaptive immune response to viral infection and oncogenesis. In a previous study focused on diffuse large B cell lymphoma^162^, we incidentally observed that, within the UK BioBank cohort of 502,506 individuals^163^, cHL is one of the cancer types most strongly associated with germline HLA-I homozygosity (156/562 of cases, 28%; p=0.005 [binomial test] versus 21% of the normal population from UK), potentially suggesting a decreased ability to present antigens (including viral ones) as a risk factor for this cancer. Other virus-associated tumors did not show a significantly high rate of HLA-I homozygosity in the UK BioBank (**Figure S11**).

Because the tumor virus status is not available in the UK BioBank, we then set out to investigate the role of germline allele type of the three major HLA-I genes (*HLA-A*, *HLA-B*, and *HLA-C*, encoding the heavy chain subunit of the MHC-I) in 8 virus-associated cancer types by performing germline HLA-I typing in a total of 1,255 patients annotated for tumor virus status. This analysis revealed that EBV-positive cHL had the highest rate of germline homozygosity in at least one HLA-I gene (8/15 cases, 53%), which was significantly higher than expected based on the rate in the general population (21%) estimated from a subset of the GTeX database^164^ (**Figure 6A**). EBV-positive cHL had a higher rate of homozygosity in *HLA-C* individually compared to EBV-negative cases (p=0.0039, Fisher’s exact test), and a similar trend was seen for *HLA-B* (p=0.17) (**Figure S10, Table S14**). While no individual HLA-I allele was significantly enriched in the small number of EBV-positive cases available, the HLA-A*02*01 allele was more frequent in the EBV-negative than EBV-positive cases (32% [13/41] versus 0%, p=0.012), consistent with what has been observed in larger population studies of HLA-I allele type in cHL^165^ (**Figure 6C**). These findings suggest germline HLA-I homozygosity may be an inherited risk factor for the development of EBV-positive cHL, possibly due to a reduced diversity of MHC molecules available for viral antigen presentation.

**Figure 6:**
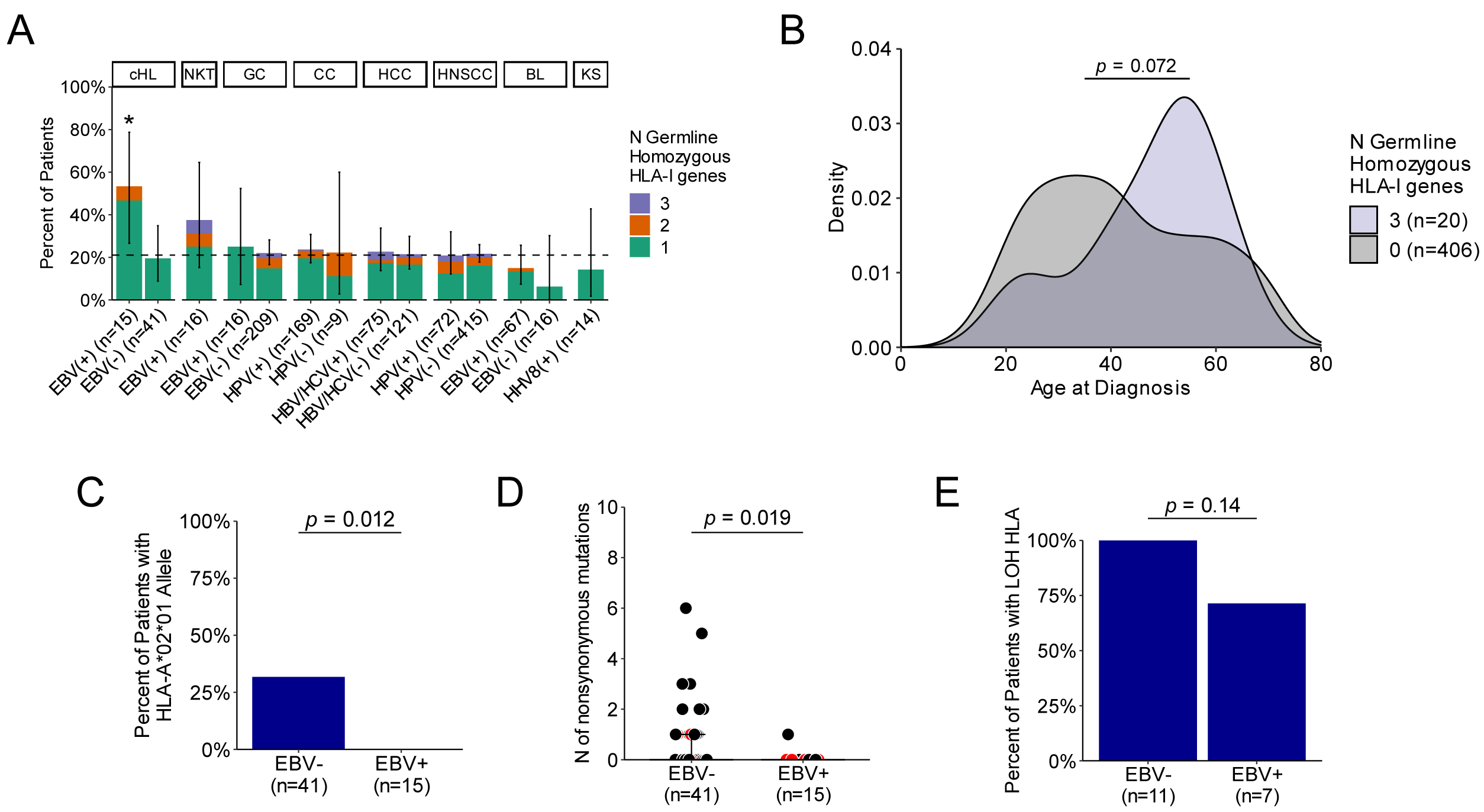
Germline and somatic status of HLA-I in EBV-negative and EBV-positive cHL. A) Percent of germline homozygous individuals in HLA-I in seven virus-associated cancers by virus-infection status (positive versus negative). * p < 0.05, binomial test. B) Distribution of age of cHL diagnosis in UK BioBank cHL patients that were germline heterozygous in HLA-I (n=406) versus germline homozygous in all three HLA-I genes (n=20). C) Percent of patients with the HLA-A*02*01 allele by virus status (n=56). P value: Fisher’s exact test. D) Count of nonsynonymous mutations in HLA-I genes in cHL by virus status (n=56). P value: MWU test. E) Percent of patients with loss of heterozygosity of HLA-I by virus status (n=18). P value: Fisher’s exact test.

Of note, cHL patients with germline homozygosity in HLA-I displayed unique phenotypic characteristics compared to those that are germline heterozygous. Specifically, they were mostly male (9/16 [56%] patients in our WES-sequenced cohort; versus 16/40 [40%], in germline heterozygous patients) and older than patients heterozygous for HLA-I (median age 52.5 in homozygous, 28.5 in heterozygous; p=0.074, MWU test). Within the UK BioBank, cHL patients who were fully heterozygous exhibited a bimodal diagnosis age curve with a larger peak from age 20-30 and a smaller peak at age 60. In contrast, cHL patients who were fully homozygous in all three HLA-I loci exhibited a diagnosis age curve with the greatest peak at ages 50-60, following a different age distribution compared to heterozygotes (p=0.072, Kolmogorov-Smirnov [KS] test) (**Figure 6B**). These patterns are consistent with the known enrichment of EBV-positive cases among older versus younger cHL patients in Western countries and may reflect a higher frequency of germline HLA-homozygous cases within the EBV-positive subtype in the UK BioBank cohort.

Next, we assessed the distribution of somatic variation at HLA-I and *B2M*, which encodes the beta-2 microglobulin subunit of the MHC-I complex, in our cohort by EBV infection status. Missense or truncating mutations in one or more HLA-I loci were detected in 17/56 cases, including 8/56 in *HLA-A*, 11/56 in *HLA-B*, and 5/56 in *HLA-C* (**Figure 3A** and **Table S15**). For all HLA-I loci, the majority of patients with missense or truncating mutations were EBV-negative: 8/8 for *HLA-A* (p=0.093), 10/11 for *HLA-B* (p=0.25), and 6/6 for *HLA-C* (p=0.31, Fisher’s exact test), collectively reaching statistical significance despite the small number of cases (n=16/41 EBV-cases, 39%, versus 1/15 EBV-positive cases, 7%; p=0.023). 2/56 cases (both EBV-negative) had biallelic nonsynonymous mutations in at least one *HLA-I* gene, and 1 case had biallelic truncating mutations in *HLA-B*. An additional 12/56 cases harbored missense or truncating mutations in *B2M* (detected through WES and/or Sanger sequencing^128^), and 3/56 cases (3/3 EBV-negative) harbored a missense or truncating mutation in both *B2M* and at least one HLA-I gene. In total, 26/56 (46%) cases were mutated in HLA-I or *B2M*, the majority of which (23/26, 88%) were EBV-negative (p=0.032; Fig. 3A). The count of nonsynonymous mutations in HLA-I per patient was greater in EBV-negative than EBV-positive cases (**Figure 6D**), consistent with the trend for greater somatic mutation load in EBV-negative cases seen genome-wide (**Figure 2, Figure S4**). Similarly, somatic loss of heterozygosity caused by allele deletion tended to occur more often in EBV-negative than EBV-positive cases (11/11 [100%] versus 5/7 [71%] evaluable cases, p=0.14, MWU test) (**Figure 6E**). Overall, EBV-negative cHL more frequently carried somatic lesions potentially disturbing MHC-I presentation of tumor neo-antigens, which in turn are likely more numerous in this group than in EBV-positive cHL, due to higher somatic mutation burden (analogous to carcinogen induced-tumors such as lung carcinoma and melanoma)^32^.

### Virus-associated cancers exhibit more frequent responses to immunotherapy

*PD-L1* overexpression has been linked to better overall survival in patients treated with immune checkpoint inhibitors (ICI) in several tumor types, including gastric cancer^166^, head and neck cancers^167^, and Merkel cell carcinomas^168^. *PD-L1* expression has been associated with infection by oncoviruses including EBV^169^, HPV^170^, HBV^171^, and MCPyV^168^. To determine whether virus positivity might be a useful marker for response to ICI therapy, we evaluated the correlation of viral status with response to ICI therapy with anti-PD(L)1 in 39 studies reported on ClinicalTrials.gov that had available therapy response and virus infection status data, representing four virus-linked cancers (**Table S16**).

Virus positivity was significantly associated with ICI therapy response in GC (OR 2.27, 95% CI 1.17-4.29, p=0.011, Fisher’s exact test) and HNSCC (OR 1.89, 95% CI 1.27-2.82, p=0.0012), but not MCC (OR 1.09, 95% CI 0.49-2.45, p=0.85) or HCC (OR 1.27, 95% CI 0.94-1.73, p=0.12) (**Figure 7A**). The same tumors displayed significant association between *PD-L1* expression and ICI therapy response, including GC (OR 3.85, 95% CI 2.29-6.72, p<1.0e-5), HCC (OR 1.52, 95% CI 1.08-2.13, p=0.012), and HNSCC (OR 1.89, 95% CI 1.01-3.80, p=0.045), whereas a trend was observed in MCC (OR 2.25, 95% CI 0.76-7.63, p=0.15). As expected, higher tumor mutational burden (TMB) was associated with ICI therapy response in both GC (OR 3.55, 95% CI 2.09-6.08, p=7.74e-7) and HNSCC (OR 4.31, 95% CI 3.25-5.71, p<2.2e-16), the only two cancer types with such data available.

**Figure 7:**
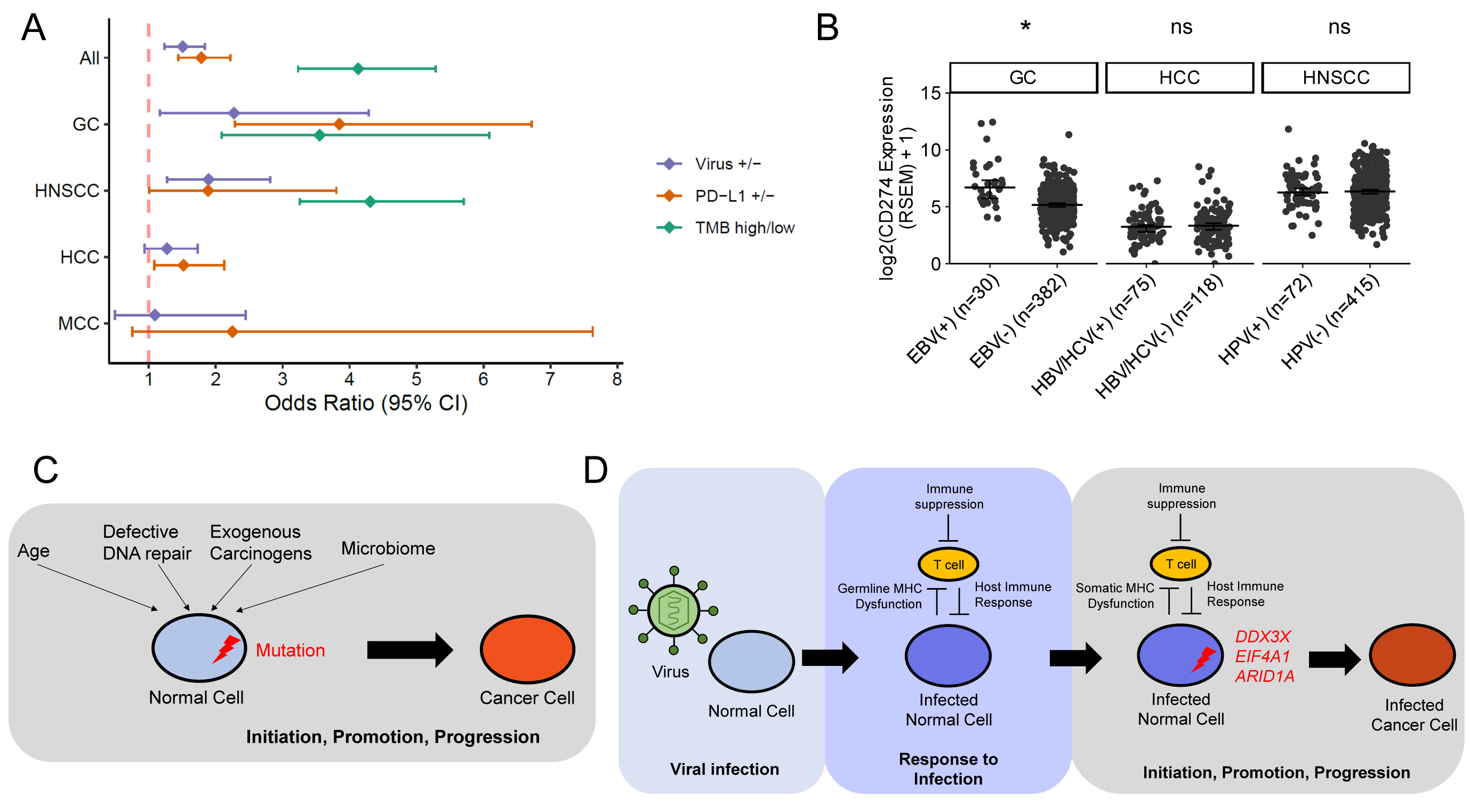
Analysis of immunotherapy trials in virus-associated cancers. A) Odds ratio of positive response to treatment with ICIs with virus-positive status, PD-L1 positive status, and/or high tumor mutation burden (TMB) in 37 studies representing four types of cancer. B) Expression of PD-L1 (CD274) versus viral status of tumors in TCGA studies of GC (TCGA-STAD), HCC (TCGA-LIHC), and HNSCC (TCGA-HNSC). * p < 0.05, MWU test. C) Model for oncogenesis in the absence of viral infection. A normal cell accumulates driver mutations as a result of age, defective DNA repair, exogenous carcinogens, or microbiome interactions, leading under selective pressure to initiation, promotion, and progression that ends in the malignant transformation of the cell. D) Model for oncogenesis in the presence of viral infection. A normal cell is infected with a virus and a latent infection is established as a result of inadequate host immune response, potentially associated with germline MHC dysfunction or other inherited risk factors. The infected normal cell acquires somatic mutations in specific genes, such as chromatin modifiers like *ARID1A* or RNA helicases *DDX3X* and *EIF4A1*, leading to initiation, promotion, and progression that ends in the malignant transformation of the infected cell.

In order to determine whether virus positivity is an independent marker of ICI therapy response, we compared the expression of *PD-L1* [*CD274*] in TCGA’s studies of GC^131^, HCC^132^, and HNSCC^134^. *CD274* expression was higher in virus-positive GC compared to virus-negative GC (median 103.03 and 34.81, p=1.3e-7), but this was not the case in HCC or HNSCC (**Figure 7B**). This is independent from TMB, as virus-positive GC and HNSCC have fewer mutations than virus-negative GC and HNSCC, respectively (Figure 2B). These results suggest that EBV-positive status could be a positive prognostic marker for patients undergoing ICI therapy for gastric and head and neck cancers associated with EBV infection, which may be correlated with *PD-L1* expression in GC but may represent an independent marker in HNSCC.

## Discussion

This study provides insights into the epidemiological, inherited, somatic, and immune components commonly implicated in the pathogenesis of cancers associated with oncoviruses. Through analysis of cancer incidence rates reported in a selection of published studies, we noted virus-associated cancers display greater incidence in males compared to females relative to non-virus-associated cancers. The greater incidence of virus-associated cancers in males may be caused in part by immunologic predisposition towards viral infection compared to females. For example, males infected with HBV are more likely to become viral carriers, whereas females infected with HBV are more likely to develop antibodies indicative of recovery and immunity to the virus^172^. In general, females have a more robust immune response to infection than males, which has been attributed to X-chromosome inactivation and regulation of the immune response by genetic, hormonal, and environmental mediators^173,174^.

Through a large-scale analysis of DNA sequencing data from 1,658 tumors collected from different studies, we found that virus-positive tumors generally display a lower mutation load compared to virus-negative tumors. It has been hypothesized that the oncogenic activity of virus-encoded proteins removes selective pressure for somatic mutations. For example, in cHL, the rarity of mutations in the PI3K-AKT signaling inhibitor *GNA13* in EBV-positive cases may be explained by the activity of LMP2a, which has been shown to activate the PI3K/AKT pathway^175^. However, unlike the other virus-associated cancers, virus-positive hepatocellular carcinomas and BLs (comprised mostly but not exclusively of eBL) have a greater mutation load compared to virus-negative cases. In HCC, this may reflect increased genomic instability of virus-positive HCC tumors resulting from integration of HBV into the host cell genome^176,177^, and/or the activity of HCV oncoproteins that inhibit DNA repair and induce double-stranded breaks in the DNA^178^. The greater genome-wide mutation load in EBV-associated BL has been attributed, at least in part, to the presence of mismatch repair and *AICDA*-mediated ASHM signatures in these cases, though the mechanism of mutagenesis by EBV in BL is yet to be elucidated^38^. Yet, EBV-positive BL had a lower driver mutation load compared to EBV-negative BL, which may be attributed to the activity of viral proteins such as EBNA1 that reduce selective pressure for drivers genetic lesion seen commonly in virus-negative Burkitt lymphomas^38^. Interestingly, we found evidence for decreased ASHM in EBV-positive cHL compared to EBV-negative cHL, showing the opposite trend from that seen in BL. This finding highlights how mutation processes may differ even in cancers associated with the same virus, potentially related to the translation of distinct viral oncoproteins from different viral latency programs.

We found that somatic mutation of the RNA helicase protein *DDX3X* was more frequent in virus-positive tumors compared to virus-negative tumors overall and for a variety of individual cancer types. *DDX3X* is a member of the DEAD (Asp-Glu-Ala-Asp) box protein family involved in multiple functions related to RNA metabolism, including transcription regulation, splicing, RNA export, and translation initiation^160^. *DDX3X* additionally functions as a component of the innate immune signaling pathway and is known to inhibit replication of viruses such as HBV by activating production of IFN-beta^160,179^. Some RNA viruses, including HCV and HIV, exploit functions of *DDX3X* to aid in viral replication^160,179^. In cancer, *DDX3X* has been described as both a tumor suppressor and an oncogene in different cancer types and even among different tumors of the same cancer type^180^. *DDX3X* is expressed in many tissues of the body and escapes X chromosome inactivation^160,180^. The relatively high frequency of mutations in *DDX3X* in virus-positive tumors and the near-exclusive male bias for truncating mutations suggests loss of function of *DDX3X* may contribute to the pathogenesis of some virus-associated cancers, particularly BL, which had the highest frequency of *DDX3X* mutations in this study and for which similar findings were recently reported in another study^161^. It is worth noticing that *DDX3X* mutations although enriched did not occur exclusively in EBV-positive BLs. Gong and colleagues reported^154^ the role of *DDX3X* and its Y-chromosome paralog *DDX3Y* in facilitating MYC-driven lymphomagenesis. Given the pleiotropic role of *DDX3X* in viral recognition and RNA processing and its higher mutational frequency in viral related tumors beyond MYC-driven lymphomas, it will be interesting to molecularly dissect the dual role of these mutations in viral and cell processes leading to tumor development.

The greater frequency of *TP53* mutations in virus-negative compared to virus-positive tumors found in this analysis may reflect how viral oncoproteins inhibit the activity of tumor suppressors. EBV-encoded oncoproteins have been shown to inhibit tumor suppressive functions of p53 and other proteins in the p53 family^181^. Similar functions have been observed in other herpesviruses, including HHV8 (associated with KS)^181^. The E6 and E7 proteins encoded by HPV inhibit p53 and Rb, respectively^31^. In these and other cancers, the lower frequency of *TP53* mutations may reflect the lack of selective pressure for *TP53* mutations due to the disruption of p53 functions by viral oncoproteins.

The results of our study indicate the immune response plays a critical role in the risk, development, and/or response to therapy of virus-positive tumors. Alterations in MHC-I were found to follow a trend in cHL specifically, where somatic inactivating events were more frequent in virus-negative compared to virus-positive tumors, possibly to limit the presentation of neo-epitopes generated by the higher mutation burden in EBV-negative cHL, whereas germline homozygosity of HLA-I was more frequent in EBV-positive cHL, possibly to limit the presentation of viral antigens. Consistent with this finding, cHL had an elevated rate of germline HLA-I homozygosity compared to normal individuals in the UK BioBank, and cHL cases homozygous in all three HLA-I loci tended to be older patients, a group known to be enriched for EBV-positivity compared to young adult cHL. More frequent germline HLA-I homozygosity in EBV-positive cHL as a means to reduce viral antigen presentation, and more frequent somatic HLA-I inactivation in EBV-negative cHL as a means to reduce tumor antigen presentation, may contribute to explain the observations that normal HLA-I surface expression by cHL tumor cells is largely preserved in EBV-positive cases and largely lost in EBV-negative cases^182^, respectively. It remains to be explained why germline HLA-I homozygosity was not increased in other EBV-associated cancers such as EBV-positive BL^162^, and why germline homozygosity represents a common feature of DLBCL, a largely non-virus-associated B-cell lymphoma^183^. One explanation may be that BL has a more restricted expression of viral latency antigens compared to cHL^33^. In DLBCL, the high rate of germline HLA-I homozygosity (26%, compared to 28% in HL and 23% in normal) was thought to contribute to limit neoantigen repertoire presentation, given the substantial nonsynonymous mutation burden typical of this disease, including ASHM^162^. Although the mutational burden of EBV-negative cHL may appear higher than DLBCL overall^129^, the phenotypic and molecular heterogeneity of the latter warrants further analyses focused on individual subtypes in larger number of cases. Future work should also verify how germline HLA-I zygosity can predispose to the development of EBV-positive cHL.

Analysis of ICI clinical trials reveals that virus-positive status could represent a positive biomarker for ICI therapy response in GC and HNSCC. The improved response to immunotherapy of EBV-positive GC patients compared to EBV-negative GC patients is hypothesized to be due to increased expression of *PD-L1*, potentially through activation of the NF-κB pathway by viral protein LMP2A^184^. This is consistent with the association between *PD-L1* expression and EBV-positive status in TCGA’s study of GC, as well as other studies that reported similar results in GC tumors^169^. The association between HPV infection and *PD-L1* expression is less clear: some studies report a link between HPV status and *PD-L1* expression^185,186^, while others find no association^187,188^, the latter of which is consistent with the results from the TCGA HNSCC dataset. It has been hypothesized that the improved response of HPV-positive HNSCC tumors to ICI therapies may be due to increased abundance of tumor infiltrating lymphocytes and CD8+ T cells in the microenvironment of virus-positive tumors^189^.

Our integrative analysis of the epidemiological and genetic factors related to virus-associated cancers highlights two approaches to oncogenesis, in the presence and absence of viral infection. In the absence of viral infection, a normal cell acquires somatic drivers towards malignant transformation through random mutations with age, defective DNA repair, exogenous carcinogens, and interactions with the microbiome, together with selection. Following the acquisition of an average of 5-7 driver mutations^190^, the normal cell transforms into a cancer cell (**Figure 7C**). In virus-positive cancers, a normal cell is first infected with a virus, potentially as a result of inherited or environmental risk factors. The infected cell acquires somatic mutations over time, though fewer are generally required compared to the non-viral scenario due to the activities of viral oncoproteins. Finally, after acquiring the requisite number of drivers, the infected cell transforms into a cancer cell, and continues to follow a trajectory of progression distinct from the non-viral scenario (**Figure 7D**). Further studies will be needed in order to understand how differences in the development and progression of tumors due to viral infection following this model may be incorporated into the development of targeted therapies.

## Data Availability

DNA sequencing data of Hodgkin lymphoma and Kaposi sarcoma samples will be deposited in the database of Genotypes and Phenotypes (dbGaP) and available upon publication. Any additional information required to reanalyze the data reported in this paper is available from the lead contact upon request.

## Acknowledgements

This work was funded by the National Institutes of Health, National Cancer Institute grants R35CA253126 (R.R.), Fondazione AIRC (Investigator Grant no. 23732 to ET) and SU2C Convergence Program (K.G., Y.N., J.B.R., and R.R.).

## Author Contributions

Conceptualization, K.G., E.T., R.R.; Methodology, K.G., R.R.; Investigation, K.G., G.S., Y.N., J.R, C.K., M.A.; Formal Analysis, K.G., Y.N., J.R.; Provision of critical samples: C.v.N., B.F.; Writing – Original Draft, K.G.; Writing – Review & Editing, K.G., L.P., E.T., R.R.; Resources, G.S., G.P., A.C., E.T., R.R.; Supervision, E.T., R.R.

## STAR Methods

### RESOURCE AVAILABILITY

#### Lead contact

Further information and requests for resources should be directed to and will be fulfilled by the Lead Contact, Raul Rabadan.

#### Materials availability

This study did not generate new unique reagents.

#### Data and code availability

- DNA sequencing data of Hodgkin lymphoma and Kaposi sarcoma samples will be deposited in the database of Genotypes and Phenotypes (dbGaP) and available upon publication.
- This paper does not report original code.
- Any additional information required to reanalyze the data reported in this paper is available from the lead contact upon request.

### EXPERIMENTAL MODEL AND SUBJECT DETAILS

#### Overview

Clinical and genomic data of 1,974 cancer patients was obtained from 14 published studies^129–131,133–137,141,149,156–159^ (1,963 patients) and newly collected DNA sequencing data of Hodgkin lymphoma (32 patients) and Kaposi sarcoma (10 patients). The combined cohort consists of 612 females, 903 males, and 356 individuals of unknown or unreported sex. The ages range from 1 year to 90 years. IDs of the newly sequenced samples are anonymized. Detailed information is provided in **Table S1**.

#### Hodgkin lymphoma

32 Hodgkin lymphoma cases were newly analyzed at the University of Perugia, Italy, as further detailed below. Additional information is provided in **Table S1**.

#### Kaposi sarcoma

10 Kaposi sarcoma patients were enrolled for study at the University of Sassari, Italy. Skin lesion tumor samples and adjacent non-neoplastic cells were surgically resected for sequencing. All biological samples (tissue and blood specimens) were obtained after the written consent from the patients. The study was approved by the Committee for the Ethics of the Research and Bioethics of the National Research Council (CNR n.12629). Additional information is provided in **Table S1**.

### METHOD DETAILS

#### Epidemiological Analysis

Sex ratio in incidence rates of virus-associated and non-virus associated cancers, as well as virus-positive and virus-negative cases of virus-associated cancers, were obtained from studies listed in **Table S2**^40-121^. Global age-standardized incidence rates of cancers by country in 2020 were obtained from GLOBOCAN 2020 Cancer Today online portal (https://gco.iarc.fr/today/home). Attributable fraction of cancer cases for each region were obtained from de Martel et al^3^.

#### Sample preparation and sequencing

##### Hodgkin lymphoma

Whole-genome sequencing (WGS) was performed on tumor and normal samples from a cohort of 27 cHL cases previously subjected to whole-exome sequencing (WES)^128^, as well as from 5 newly microdissected EBV+ cHL cases, under an IRB-approved protocol after written informed consent^128^. For each patient, we laser-microdissected HRS cells (n=1200-1800 per case) along with a similar number of adjacent nonneoplastic cells from frozen lymph node sections and subjected the samples to whole genome amplification (WGA) in duplicate. Here, whole genome sequencing (WGS) was performed separately for each tumor duplicate at a median depth of 44X, as well as for the pooled normal duplicates to a median depth of 44X (**Table S4**). For 5 cases previously subjected to WES^128^, we additionally sequenced the whole genome of unamplified DNA from peripheral blood at a median depth of 41X. Preparation of libraries for WGS was done using Illumina TruSeq DNA PCR-Free library kit and Illumina TruSeq Nano DNA library kit for 31/32 cases and 1/32 case respectively, followed by paired-end sequencing for 2×125 cycles on Illumina HiSeq 2500 and for 2×150 cycles on Illumina NovaSeq instruments for 20/32 cases and 12/32 cases, respectively.

#### Virus infection status calling

EBV infection status of cHL patients was determined by standard EBER in-situ hybridization on fixed tissue sections and/or confirmed presence of reads aligned to the EBV reference assessed by samtools idxstats. Virus-infection status of patients in other cohorts were reported in the original studies^38,128–130,132,135–137,156–159^ or obtained via cBioportal (https://www.cbioportal.org/) for TCGA cervical^133^, gastric^131^, and head and neck squamous cell carcinoma^134^ data sets.

#### Single nucleotide and indel variant calling pipeline

##### Hodgkin lymphoma

Whole genome sequencing samples were aligned to GRCh37 using the Burrows-Wheeler aligner. Samples were pre-processed by indel realignment, duplicate removal, and base recalibration with GATK^191^ following the GATK best practices workflow^192^. SAVI-v2^193^, an in-house variant caller, was used to call somatic variants. As there were two microdissected tumor and one normal sample for each case sequenced by WGS, we adapted our previously described WES pipeline^128^ for one normal microdissected sample rather than two by defining somatic variants present in a major tumor clone as those having a VAF >= 20% in both tumor replicates, <3% in the pooled normal microdissected sample, and <1% in the unamplified blood sample (when available) at any genome bases that were covered by at least 6 reads in all tumor, normal and blood samples of each case (representing a median of 89% of all genome bases, IQR 87-90%). The threshold of 6 reads was selected because, in a comparison to deeper WES data from the same cases (n=31 with at least 1 mutation called on both WES and WGS) taken as benchmark (median coverage 148X), this threshold provided absolute specificity (99.999%) while preventing the loss of sensitivity at minimum depths higher than 6 (**Figure S12**). All other filters, including those to remove SNPs, strand bias, and homopolymeric indels, were applied as previously described^128^. To further account for errors in the whole genome sequencing data, we removed variants within ENCODE or Duke consensus blacklist regions (http://genome.ucsc.edu/cgi-bin/hgFileUi?db=hg19&g=wgEncodeMapability), as those are suspected artifacts. Finally, we used normal samples from all 32 WGS cases to construct a cohort supernormal and removed any putative somatic variants in these cases that was called also in the supernormal.

WES data were analyzed for 38 cHL cases of ours (34 already published plus 4 newly microdissected and processed as in ref.^128^, except for using the updated Agilent SureSelectXT V6+Cosmic probes and kit), following a bioinformatics pipeline previously described^128^ that was also applied to the WES data of 18 cHL cases available for download from ref.^129^. The latter data were generated after flow cytometry sorting of tumor and normal cells from cryopreserved tissue cell suspensions, without subsequent WGA, and comprised one tumor and one normal sample per case. These 56 WES samples were subjected to the same bioinformatics pipelines for mutation calling (described in ref.^128^) and for copy number aberration calling and HLA-I analyses (described further below). Two cases (patient IDs c_cHL_4 and c_cHL_24) were excluded from mutation load comparison due to previously described microsatellite instability^129^.

##### Kaposi sarcoma

WES data from 10 samples were aligned to GRCh37 using the Burrows-Wheeler aligner. Samples were pre-processed by indel realignment, duplicate removal, and base recalibration with GATK following the GATK best practices workflow. SAVI-v2^193^ was used to call somatic variants. The variant list was filtered for variants with a minimum total depth 10 and maximum total depth 700 in both tumor and normal, strand bias p value > 0.001 in tumor and normal, and called as significant somatic variants by SAVI (p-value <0.05, and confidence interval for the significance of the tumor/normal comparison >0). Variants were excluded if they were found in an in-house supernormal created from 186 normal samples from the TCGA, if they were in the cohort supernormal constructed from variants in the ten normal samples, or if they were common SNPs found at a frequency >= 5% in the 1000 Genomes Project.

#### Mutation load analysis

The mutations in each tumor were obtained from the variant lists reported in the original studies (for previously published cases^129–131,133–137,141,149,156–159^), or from the mutation calling pipeline described above (for newly sequenced Hodgkin and Kaposi sarcoma cases). Mutations in driver genes were defined as mutations that occurred within genes described as cancer-specific drivers and/or recurrently mutated genes in the original studies^129–131,133–137,141,149,156–159^. Aberrant somatic hypermutation (ASH) mutations were defined as mutations occurring in as regions within 2 kb of the transcription start site of 126 previously identified targets of ASHM [75, 126, 127], as previously described [33]. ASH mutations per Mb were calculated by dividing the number of mutations by the total length of the candidate regions (0.252 Mb).

#### Mutation signature analysis

Mutation signatures were called from somatic variants separately for each cancer type using Palimpsest^194^, an NMF-based mutation signature caller. Signatures were obtained from somatic variants called from whole genome sequencing data when available (cHL, Burkitt) or whole exome sequencing data (other cancers). First, unsupervised mutation signature analysis was run for single base substitution (SBS), double base substitution (DBS), and insertion-deletion (ID) signature calling (when applicable) for all variants in all patients of a given tumor type. The similarity between de novo inferred signatures and published mutation signatures from COSMIC v3 was assessed by cosine similarity function in Palimpsest. Supervised mutation signature analysis was then run separately for each cancer type with mutation signatures that were the highest ranking match for the inferred signature from the unsupervised analysis.

Mutation signature calling in cHL samples followed the methods above with the addition of a filter for clonal variants only (Tumor VAF >=20% in both replicates) to account for noise in whole genome amplified samples. Additionally, to improve detection of potential AID-associated signatures expected in this data set due to the germinal center B cell of origin of this tumor, de novo signatures were also separately called for clustered variants, i.e. variants grouped by nearest mutation distance (NMD) below a threshold defined according to approach of ref.^195^, i.e. NMD < 316 bases for WGS based on visual inspection of the histogram of NMD (**Figure S13**). All de novo signatures found in cHL cases were compared to published whole genome amplification signatures^196^ to rule out sequencing artifacts from the amplification procedure. All other steps of the mutation signature analysis were performed as described above.

#### Copy number segmentation and variant calling

Copy number segmentation of cHL samples was conducted using Control-FREEC^197^ for each pair(s) of tumor and normal samples sequenced by whole genome sequencing available for each case. For patients with more than one tumor and/or normal sample, only aberrations occurring in overlapping regions of gain or loss present in both tumor replicates and in neither normal sample were counted as non-normal copy number. Copy number aberrations were defined based on the following absolute CN cut off values: CN > 2.3 gain, CN > 3.6 amplification, CN < 1.7 heterozygous loss, and CN < 0.8 homozygous loss. The CN of overlapping regions of CNA was calculated as the mean of the constituent copy numbers. When compared to fluorescence in situ hybridization data already available for JAK2 and TNFAIP3 deletion in 33/34 and 32/34 previously published cases, sensitivity and specificity of this analysis for focal JAK2 gain and TNFAIP3 loss on WGS cases were 38% and 94% for JAK2 copy number gain, and 29% and 81% for TNFAIP3 deletion (**Table S6**).

Copy number segmentation of Kaposi sarcoma samples was conducted using Sequenza^198^ for each pair of tumor of normal samples sequenced by whole exome sequencing for each case. Copy number segmentation of plasmablastic lymphoma samples was performed with Oncoscan, as previously described^130^. Copy number segmentation data of other cancers was obtained from the original published studies (Burkitt^149^, NKTCL^158^) or cBioportal (TCGA samples^131–134^).

#### GISTIC Analysis

##### Hodgkin lymphoma

In order to define significant regions of recurrent CNAs in Hodgkin lymphoma, GISTIC 2.0^199^ was applied to the copy number segmentation of 56 cases of cHL sequenced by WES with a q < 0.2, a maximum segmentation threshold of 10,000, and all other parameters default via the GenePattern server (https://www.genepattern.org/). To denoise the list of putative recurrent CNAs in the WES cases, GISTIC 2.0 was applied to the cohort of 32 cases of cHL sequenced by WGS, with a q < 0.5, a maximum segmentation threshold of 10,000, and all other parameters default via the GenePattern server. The list of putative CNA regions defined by WES was filtered to include only those peaks occurring on the same cytoband as a peak called in the WGS cohort, and/or in a region previously described as a known site of recurrent copy number aberration in B cell lymphomas by manual curation. Significant peaks were defined as those having a q* < 1e-04, with q* = sqrt(q_WES_ * q_WGS_), q_WES_ is the q value of the peak in the cytoband called by GISTIC in the WES cohort and q_WGS_ is the q value of the peak in the cytoband called by GISTIC in the WGS cohort. GISTIC peaks of gain or amplification were counted as present in a patient if the maximum inferred tumor copy number within the wide peak limit region was > 2.3 (gain) or > 3.6 (amplification). GISTIC peaks of deletion were counted as present in a patient if the minimum inferred tumor copy number within the wide peak limit region was < 1.7 (heterozygous loss) or < 0.8 (homozygous loss). Arm and whole-chromosome level CNAs were defined as lesions of the same type (i.e. gain or loss) that covered >75% of the chromosome arm or chromosome, respectively.

##### Pan-cancer

In order to define significant regions of recurrent CNAs across virus-associated cancers, GISTIC 2.0^199^ was applied to pooled copy number segmentation data of 1,557 tumors, using a significance threshold of q = 0.1, a maximum segmentation threshold of 10,000, and all other parameters default via the GenePattern server (https://www.genepattern.org/) (**Table S10**). GISTIC peaks of gain or amplification were counted as present in a patient if the maximum inferred tumor copy number within the wide peak limit region was > 2.3 (gain) or > 3.6 (amplification). GISTIC peaks of deletion were counted as present in a patient if the minimum inferred tumor copy number within the wide peak limit region was < 1.7 (heterozygous loss) or < 0.8 (homozygous loss). Arm and whole-chromosome level CNAs were defined as lesions of the same type (i.e. gain or loss) that covered >75% of the chromosome arm or chromosome, respectively.

#### Analysis of Recurrently Mutated Genes in Virus-associated Cancers

Nonsynonymous mutations in protein-coding genes were counted in 1,658 patients with DNA sequencing data. For patients with both WES and WGS available, WES was used due to greater depth of sequencing. Hodgkin lymphoma cases were filtered to include only clonal mutations (Tfreq >=20; in both tumor samples when applicable) to account for noise in cases that had undergone whole genome amplification. Cases of MCC and PCNSL were only counted in the statistics for genes which were included in those targeted panels. To eliminate noise from hypermutated samples, for all cancers, only cases with <300 mutations were included. Significant genes were defined as those with an odds ratio (OR) of mutation in virus positive versus negative >1 and BH corrected p value <0.05 and recurrently mutated in at least two cases in at least two unique cancer types. Significant copy number alterations were defined as those identified from the pan-cancer GISTIC analysis (see section “GISTIC analysis” above) with an odds ratio (OR) of mutation in virus positive versus negative >1 and BH corrected p value <0.0001.

#### GNA13 sequencing

A two-round DNA PCR amplification was performed for GNA13 coding exons (fully nested for exons 1, 2 and 4; seminested for exon 3) in 18 additional EBV+ cHL cases identified in the Perugia and Amsterdam hematopathology archives. For each sample, at least 50 tumor cells were microdissected (along with a comparable number of non-tumor cells) from frozen lymph node sections stained with hematoxylin and eosin, or with immunohistochemistry for CD30. Cell microdissection was performed with a Palm/Olympus laser microdissector as described in ref.^128^. Microdissected cells were then lysed at 95°C for 10 minutes in PCR buffer, and PCR products were gel-purified and directly Sanger-sequenced. PCR primers and amplification conditions are available upon request.

#### HLA Analysis

##### Molecular Data Sets

Class I HLA allele typing of DNA sequencing samples (56/56 cHL^141^, 10 newly sequenced Kaposi sarcoma, 83 BL^149^) was performed using PolySolver with default parameters. Class I HLA allele typing of 4 additional previously published Kaposi sarcoma samples^200^ and all NKTCL samples^158^ from RNA sequencing was performed using arcasHLA^201^ with default parameters. Class I HLA typing of TCGA data sets from RNA-seq was obtained from a previous study^202^. We counted as “homozygous” those cases where both inferred alleles of an HLA-I gene are the same to the two-field resolution (allele group and specific HLA protein). For cHL cases with multiple tumor and normal samples from the same patient (38/56 patients), alleles were called from sequenced unamplified blood samples when available. Otherwise, cases were called as homozygous only if they were called as homozygous by PolySolver in all tumor and normal samples. HLA somatic mutation calling in tumor samples was performed using PolySolver against matched normal samples. Somatic HLA loss of heterozygosity, which was feasible only in non-whole genome amplified cases (n=18), was assessed with LOHHLA with a minimum coverage threshold of 5 reads using purity and ploidy inferred by Sequenza^198^ with default parameters. Cases were called as having somatic HLA loss of heterozygosity if the p value for loss of the allele was < 0.01 and the inferred copy number of the allele was < 0.5, following thresholds used in McGranahan et al^203^. HLA loss of heterozygosity was evaluated only for patients that did not undergo whole genome amplification because allele dropout from the amplification procedure prevented accurate HLA-I loci allelic copy number calling in those samples.

##### UK BioBank

Imputed HLA-I genotypes of 488,265 patients from the UK BioBank reported by HLA*IMP:02 were obtained as described in ref.^162^. UK BioBank individuals were categorized into different cancer groups by organ location/pathology according to hospital and cancer registry records using ICD10 codes. The rate of homozygosity in the general population was estimated from RNA-seq of 95 samples representing 5 tissue types in the GTEx data base, as previously described^162^.

#### Analysis of Immunotherapy Trials

The comparative analysis of response to immunotherapy was performed using data from ClinicalTrials.gov. Eleven checkpoint inhibitors targeting either PD-1, PD-L1, or CTLA-4 were included: Ipilimumab, Nivolumab, Pembrolizumab, Cemiplimab, Atezolizumab, Avelumab, Durvalumab, Camrelizumab, Sintilimab, Toripalimab, Tremelimumab. On September 22^nd^ 2022, trials were collected using the following query: “((NOT NOTEXT) [CITATIONS]) AND (OR OR …)”, where ICI drug names correspond to the ones listed above as well as any known synonyms (e.g. Ipilimumab: BMS-734016, MDX-010, MDX-101, Yervoy). Only single-arm, interventional studies where the objective response rate was available specifically for immune checkpoint therapy (with no other combination therapies) were included. Whenever possible, response stratified by virus status, PD-L1 expression and tumor mutational burden was collected. “PD-L1 positive” refers to patients that were classified as PD-L1 positive in the original study (most often, >=1% tumor cells). “Tumor mutational burden (TMB) high” refers to patients classified as TMB high in the original study (with different cut-offs used depending on the study and/or tumor type).

### QUANTIFICATION AND STATISTICAL ANALYSIS

Analyses of significance of mutation counts and frequencies were performed using a Mann-Whitney U test and two-sided Fisher’s exact test, respectively. Significance of mutation signature activities were assessed using Student’s t test. Odds ratios of mutation by virus status were computed with Haldane-Anscombe correction when applicable. The 95% confidence interval of odds ratios is reported as the normal approximation (Wald). The distribution of HLA-I germline homozygosity in HL patients in the UK BioBank was compared using a Kolmogorov-Smirnov test. Multiple hypothesis corrections were applied using the Benjamini-Hochberg Procedure and reported as q-values.

## Supplemental Information

Document S1. Figures S1-S13

Figure S1. Geographic distributions of Kaposi sarcoma and cervical cancer by country reported by GLOBOCAN 2020. A) Estimated age-standardized incidence rate (ASR) of Kaposi sarcoma by country. B) Estimated ASR of cervical cancer by country.

Figure S2. Percent driver mutations in virus positive vs negative tumors in nine cancers. P values from MWU test.

Figure S3. Somatic mutation burden of EBV-negative versus EBV-positive cHL sequenced by whole genome sequencing. Cases in the mixed-cellularity (MC) subtype are highlighted in red. A) Count of clonal mutations in coding regions. B) Count of clonal, nonsilent mutations in coding regions. C) Percentage of total mutations that are indel mutations per case. P values from MWU test.

Figure S4. Counts of recurrent CNV lesions in cHL. A) Count of copy gains in 56 cHL sequenced by WES. B) Count of copy deletions in 56 cHL sequenced by WES. C) Count of copy gains in 32 cHL sequenced by WGS. D) Count of copy deletions in 32 cHL sequenced by WGS. P values from MWU test.

Figure S5. Counts of mutations attributed to each mutation signature in virus-positive and virus-negative cases of five cancers. A) cHL (n=32); B) MCC (n=71); C) GC (n=436); D) HNSCC (n=487); E) BL (n=91). q values from Student’s t test, BH corrected.

Figure S6. Patterns of somatic hypermutation in cHL sequenced by WGS. A) Number of ASHM-associated genes mutated in 32 cHL sequenced by WGS. B) Number of mutations in ASHM regions per ASHM-associated gene in 32 cHL sequenced by WGS. P values from MWU test.

Figure S7. Odds ratio of mutation in virus positive versus virus negative tumors by cancer type. Genes with a significant OR of mutation in virus positive tumors from the combined cohort are shown. * q < 0.05, chi-square test, BH corrected.

Figure S8. A) Odds ratio of CNA in virus positive versus virus negative tumors in the combined cohort. B) Odds ratio of CNA in virus positive versus virus negative tumors by cancer type in regions with a significant OR of CNA in virus positive tumors from the combined cohort and the top ranking region with a significant OR of CNA in virus negative tumors from the combined cohort. * q < 0.0001, chi-square test, BH corrected.

Figure S9. Expression of DDX3X and DDX3X mutation status in TCGA-HNSC (n=487).

Figure S10. HLA-I gene-specific rates of germline homozygosity in 56 cHL sequenced by WES. A) Rate of germline homozygosity in HLA-A. B) Rate of germline homozygosity in HLA-B. C) Rate of germline homozygosity in HLA-C.

Figure S11. Frequency of homozygosity in HLA-I in virus-associated cancers in the UK BioBank.

Figure S12. Accuracy, sensitivity, and specificity curves used to determine depth cut-off for variant calling of cHL cases sequenced by WGS.

Figure S13. Histogram of mutation count by nearest mutation distance (NMD) used to determine cut-off for cHL de novo mutation signature analysis.

Table S1. Clinical characteristics of patients in study.

Table S2. Studies included in Figure 1B (M/F incidence rates).

Table S3. Mean count of nonsynonymous mutations in virus-positive and virus-negative tumors in 9 cancers.

Table S4. Depth of sequencing of newly sequenced WGS cHL cases Table S5. GISTIC peaks in cHL.

Table S6. Validation of JAK2 gain / TNFAIP3 loss in cHL copy number results.

Table S7. Cosine similarities of de novo mutation signatures to COSMIC mutation signatures. Table S8. Relative and absolute activities of COSMIC mutation signatures.

Table S9. Activities of MMR signatures in gastric cancer by EBV infection status.

Table S10. GISTIC peaks in virus cohort.

Table S11. Recurrently mutated genes in virus-associated cancers compared to non-virus associated cancers (and vice versa).

Table S12. Larger Burkitt lymphoma cohort for DDX3X and EBV association analysis.

Table S13. Mutations in DDX3X and EIF4A1 in 1,974 tumors.

Table S14. HLA typing in 56 cHL.

Table S15. Nonsynonymous mutations in HLA in 56 cHL.

Table S16. Studies included in Figure 7A (biomarkers of immunotherapy response).

