## SupplementaryFigures for "Genomic landscape of virus-associated cancers"

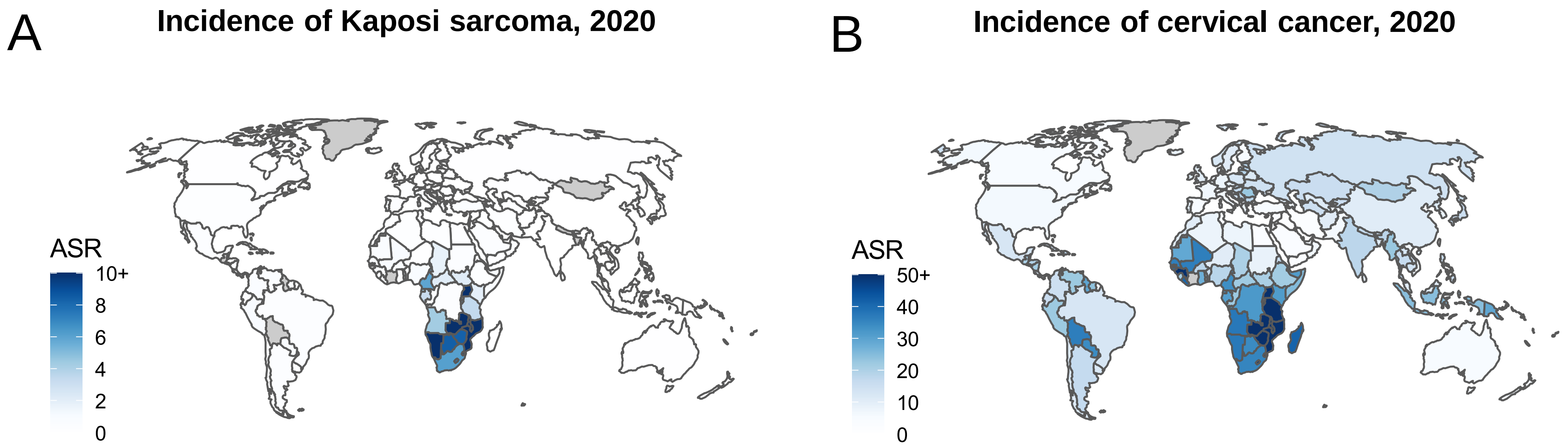
A
Incidence of Kaposi sarcoma, 2020
ASR
10+
8
6
4
2
0
B
Incidence of cervical cancer, 2020
ASR
50+
40
30
20
10
0

S1

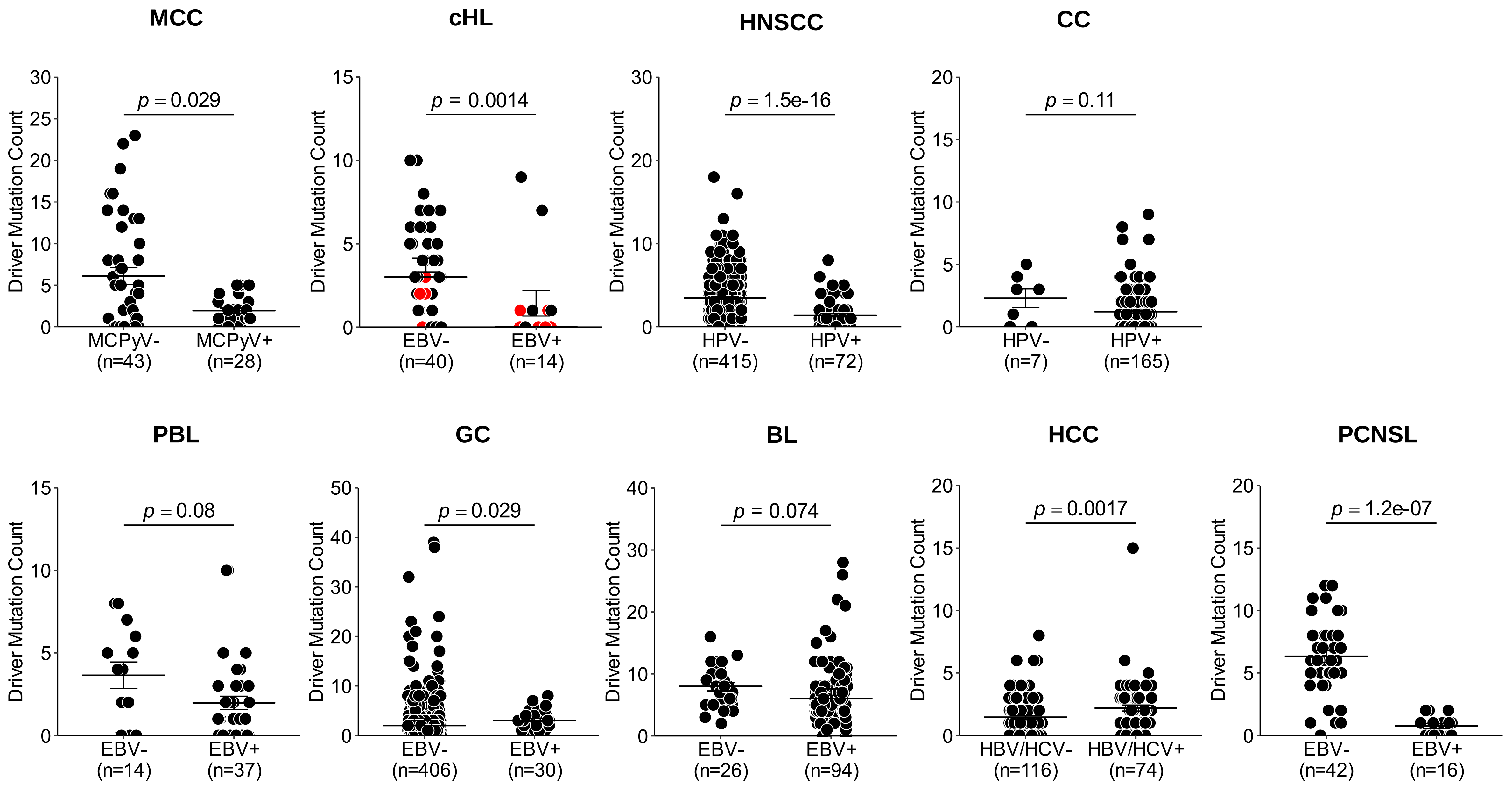
MCC
cHL
HNSCC
CC
Driver Mutation Count
p = 0.029
p = 0.0014
p = 1.5e-16
p = 0.11
MCPyV-
(n=43)
MCPyV+
(n=28)
EBV-
(n=40)
EBV+
(n=14)
HPV-
(n=415)
HPV+
(n=72)
HPV-
(n=7)
HPV+
(n=165)
PBL
GC
BL
HCC
PCNSL
p = 0.08
p = 0.029
p = 0.074
p = 0.0017
p = 1.2e-07
EBV-
(n=14)
EBV+
(n=37)
EBV-
(n=406)
EBV+
(n=30)
EBV-
(n=26)
EBV+
(n=94)
HBV/HCV-
(n=116)
HBV/HCV+
(n=74)
EBV-
(n=42)
EBV+
(n=16)

S2

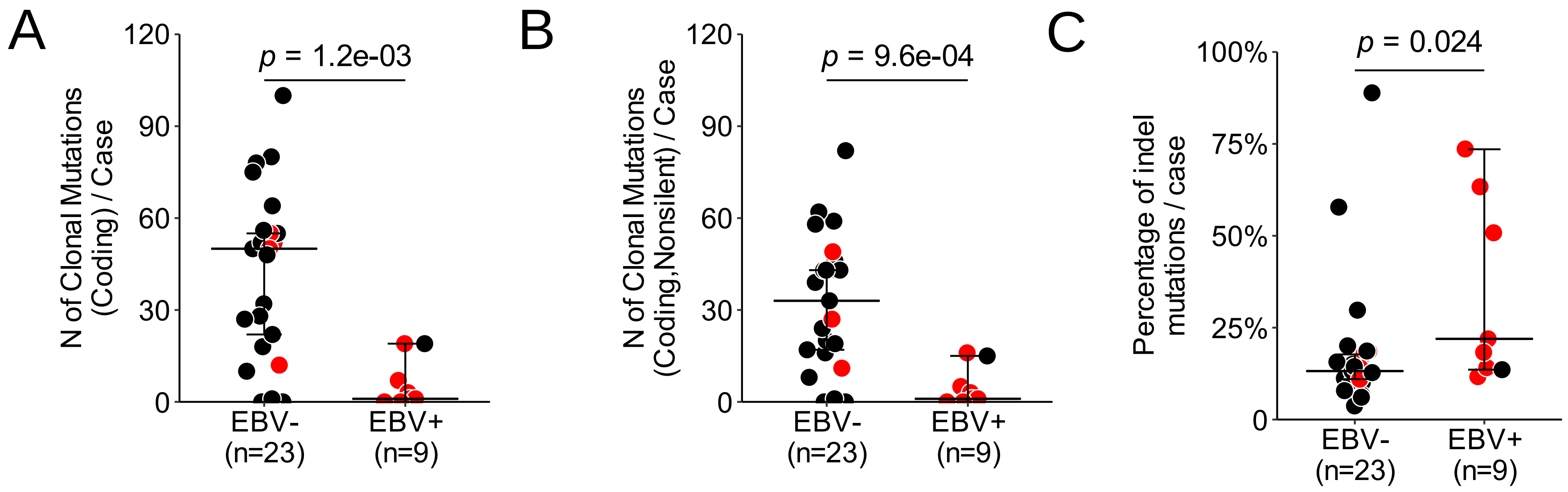
A
120
90
60
30
0
N of Clonal Mutations (Coding) / Case
p = 1.2e-03
EBV- (n=23)
EBV+ (n=9)
B
N of Clonal Mutations (Coding,Nonsilent) / Case
p = 9.6e-04
C
100%
75%
50%
25%
0
Percentage of indel mutations / case
p = 0.024

S3

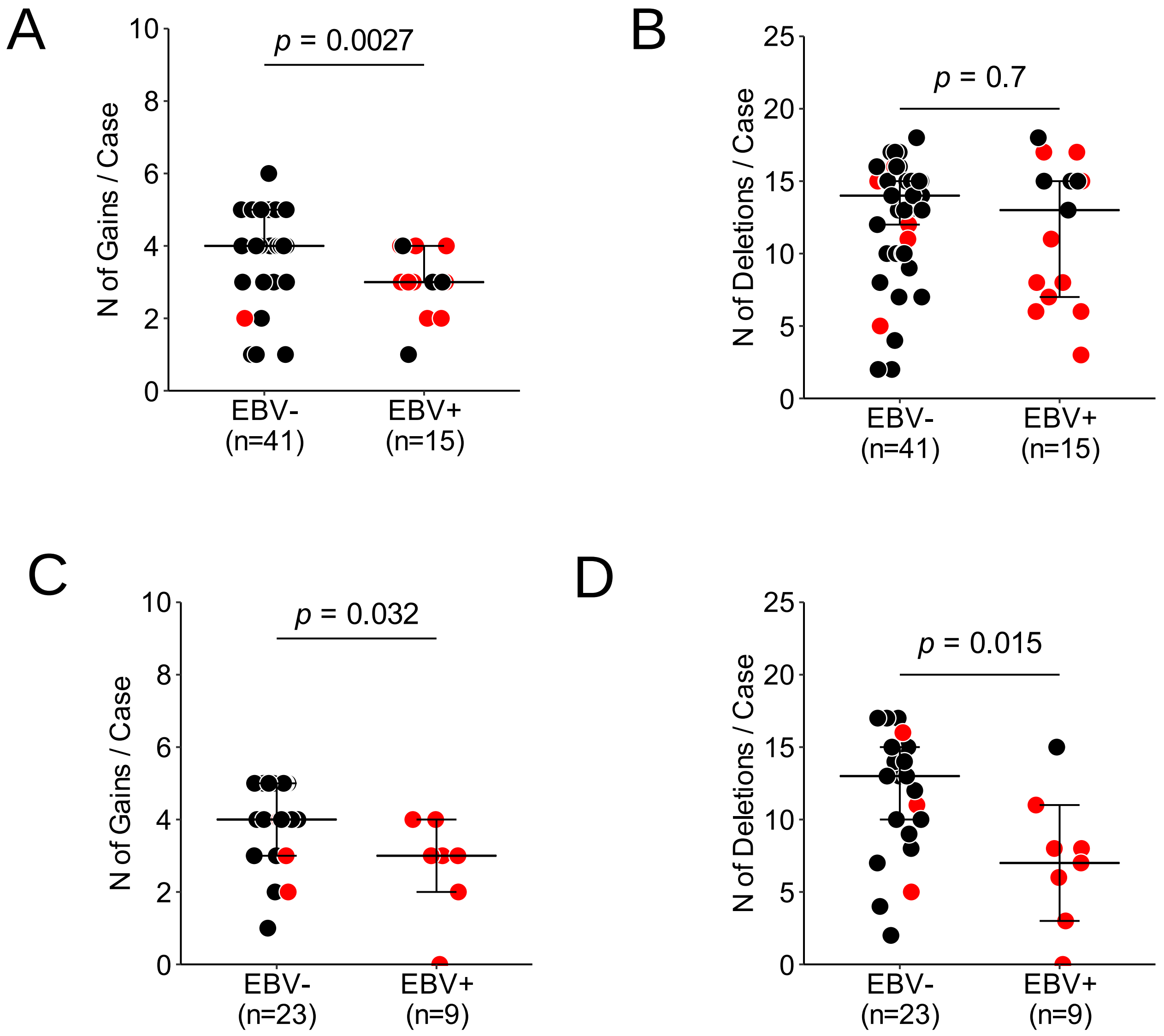
A
N of Gains / Case
p = 0.0027
EBV-
(n=41)
EBV+
(n=15)
B
N of Deletions / Case
p = 0.7
EBV-
(n=41)
EBV+
(n=15)
C
N of Gains / Case
p = 0.032
EBV-
(n=23)
EBV+
(n=9)
D
N of Deletions / Case
p = 0.015
EBV-
(n=23)
EBV+
(n=9)

S4

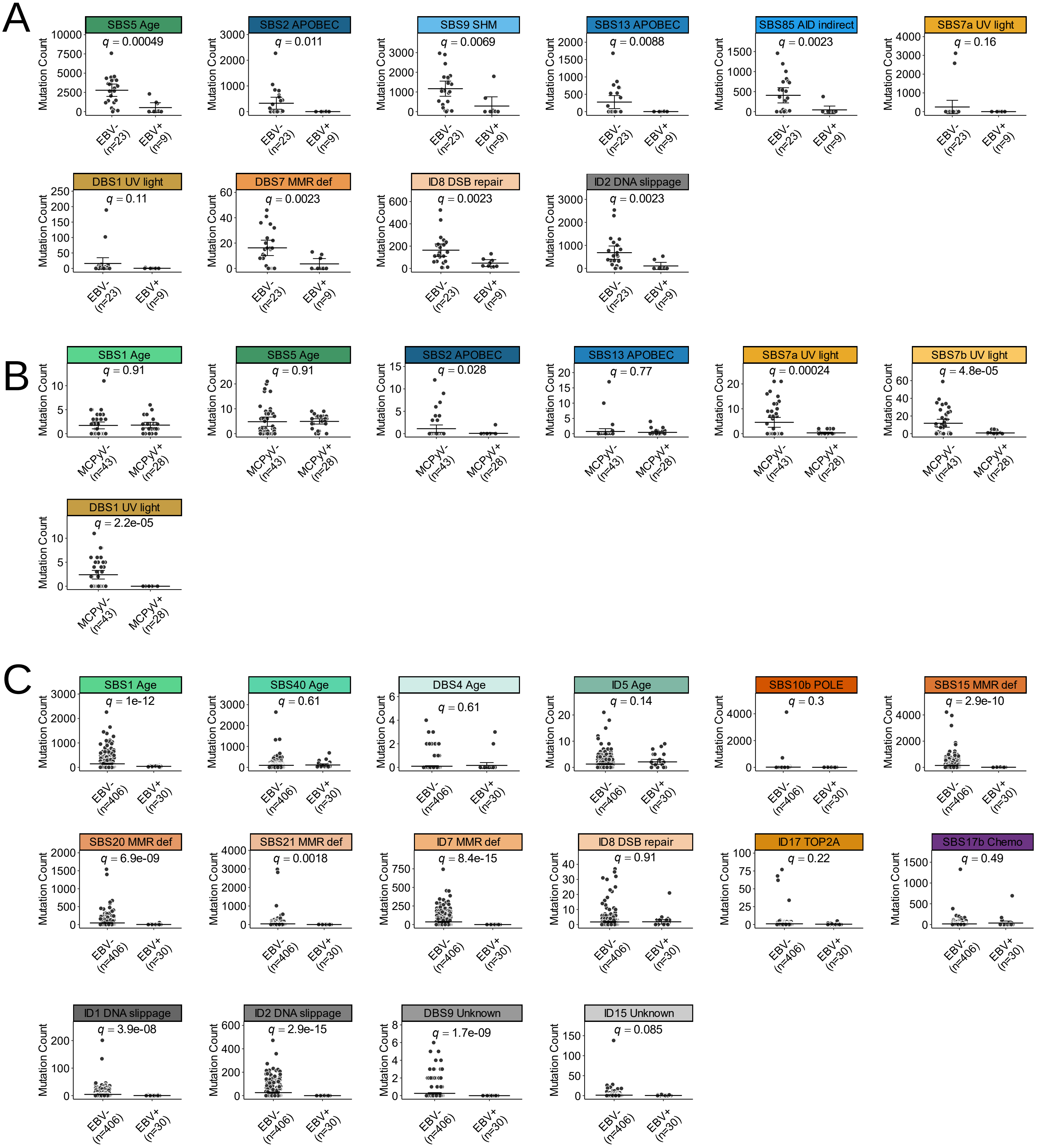
A
SBS5 Age
q = 0.00049
SBS2 APOBEC
q = 0.011
SBS9 SHM
q = 0.0069
SBS13 APOBEC
q = 0.0088
SBS85 AID indirect
q = 0.0023
SBS7a UV light
q = 0.16
DBS1 UV light
q = 0.11
DBS7 MMR def
q = 0.0023
ID8 DSB repair
q = 0.0023
ID2 DNA slippage
q = 0.0023
EBV- (n=23)
EBV+ (n=9)
Mutation Count
B
SBS1 Age
q = 0.91
SBS5 Age
q = 0.91
SBS2 APOBEC
q = 0.028
SBS13 APOBEC
q = 0.77
SBS7a UV light
q = 0.00024
SBS7b UV light
q = 4.8e-05
DBS1 UV light
q = 2.2e-05
MCPyV- (n=43)
MCPyV+ (n=28)
C
SBS1 Age
q = 1e-12
SBS40 Age
q = 0.61
DBS4 Age
q = 0.61
ID5 Age
q = 0.14
SBS10b POLE
q = 0.3
SBS15 MMR def
q = 2.9e-10
SBS20 MMR def
q = 6.9e-09
SBS21 MMR def
q = 0.0018
ID7 MMR def
q = 8.4e-15
ID8 DSB repair
q = 0.91
ID17 TOP2A
q = 0.22
SBS17b Chemo
q = 0.49
ID1 DNA slippage
q = 3.9e-08
ID2 DNA slippage
q = 2.9e-15
DBS9 Unknown
q = 1.7e-09
ID15 Unknown
q = 0.085
EBV- (n=406)
EBV+ (n=30)

S5

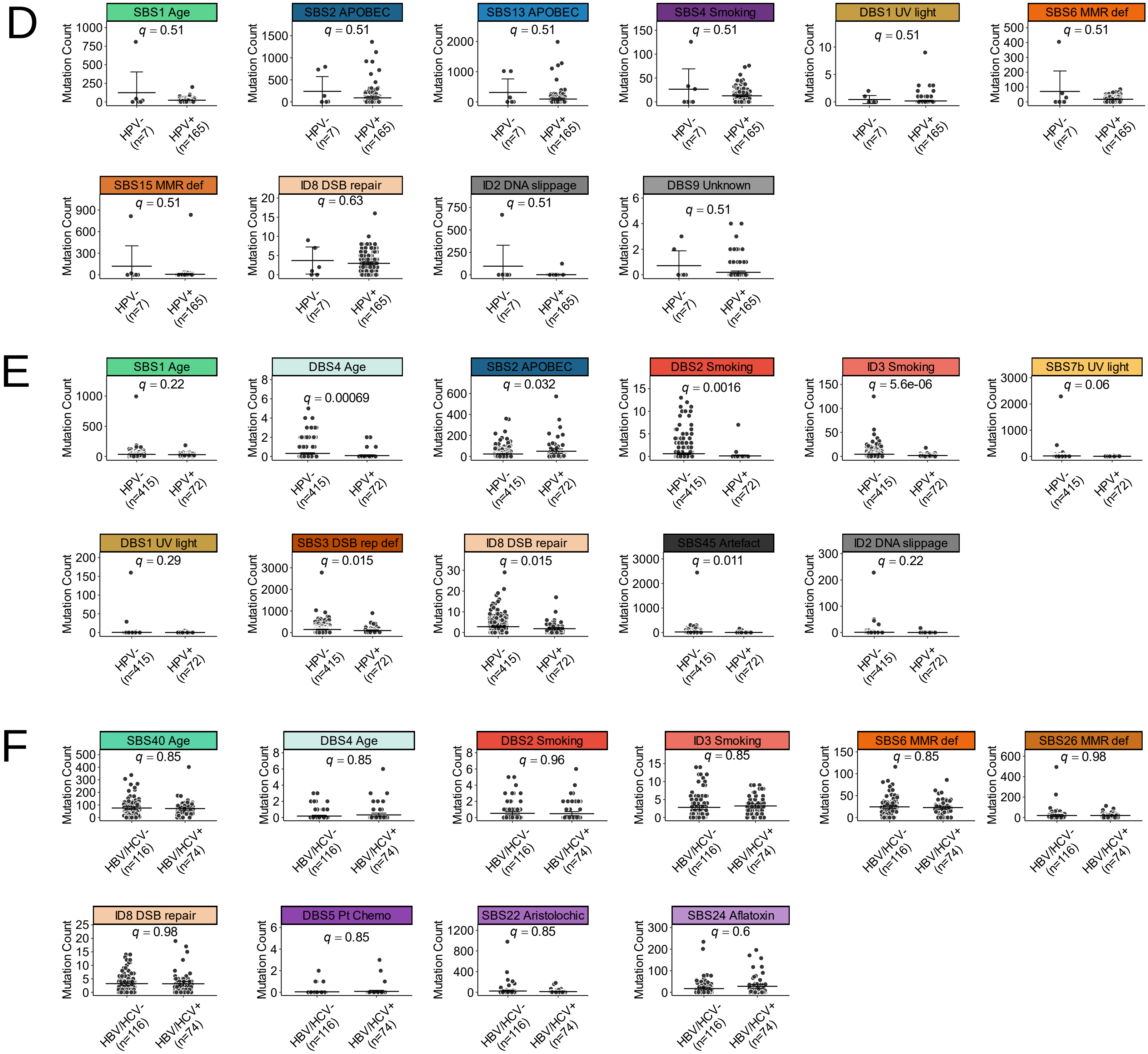


S5 (continued)

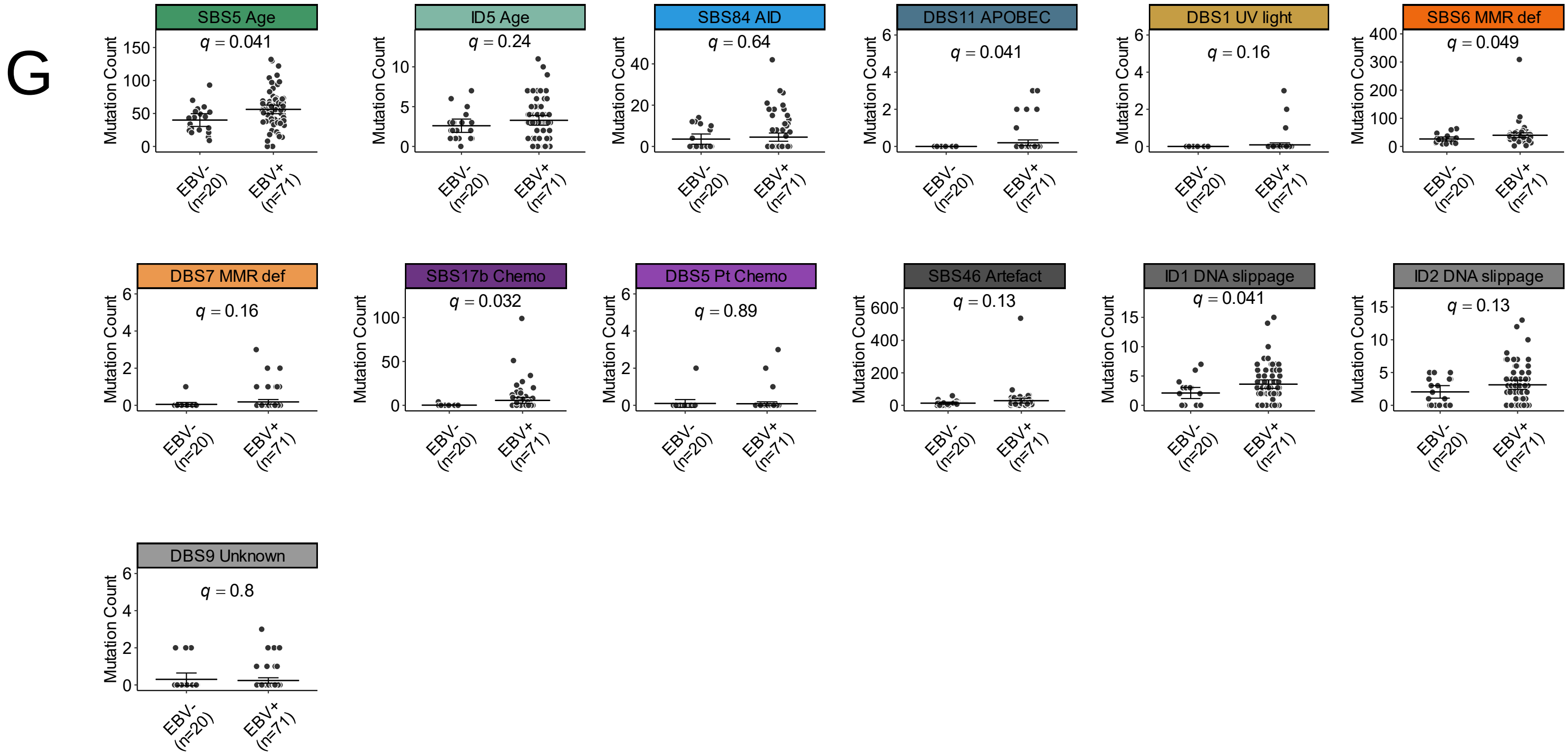


S5 (continued)

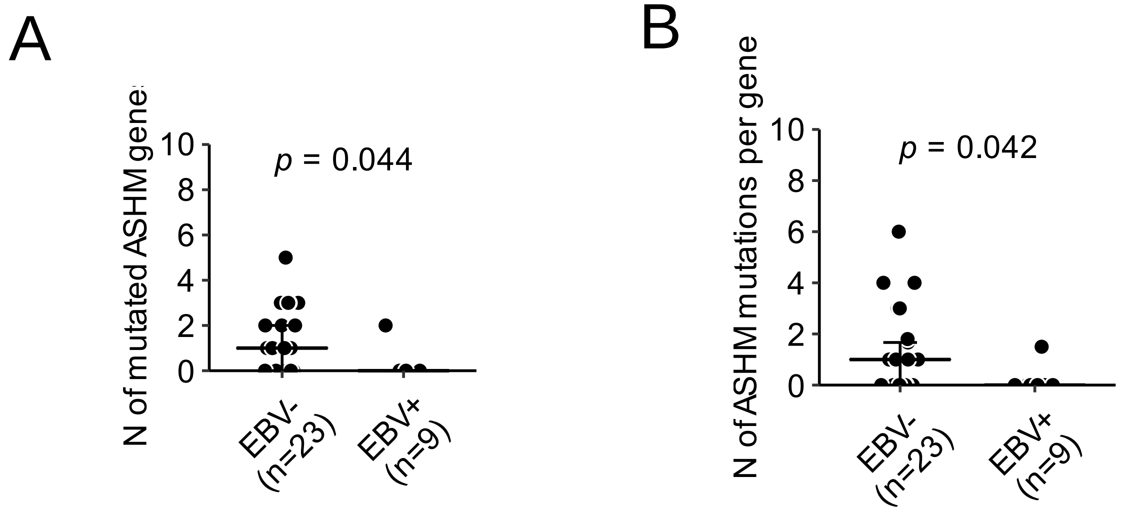
A
N of mutated ASHM genes
p = 0.044
10
8
6
4
2
0
EBV- (n=23)
EBV+ (n=9)
B
N of ASHM mutations per gene
p = 0.042
10
8
6
4
2
0
EBV- (n=23)
EBV+ (n=9)

S6

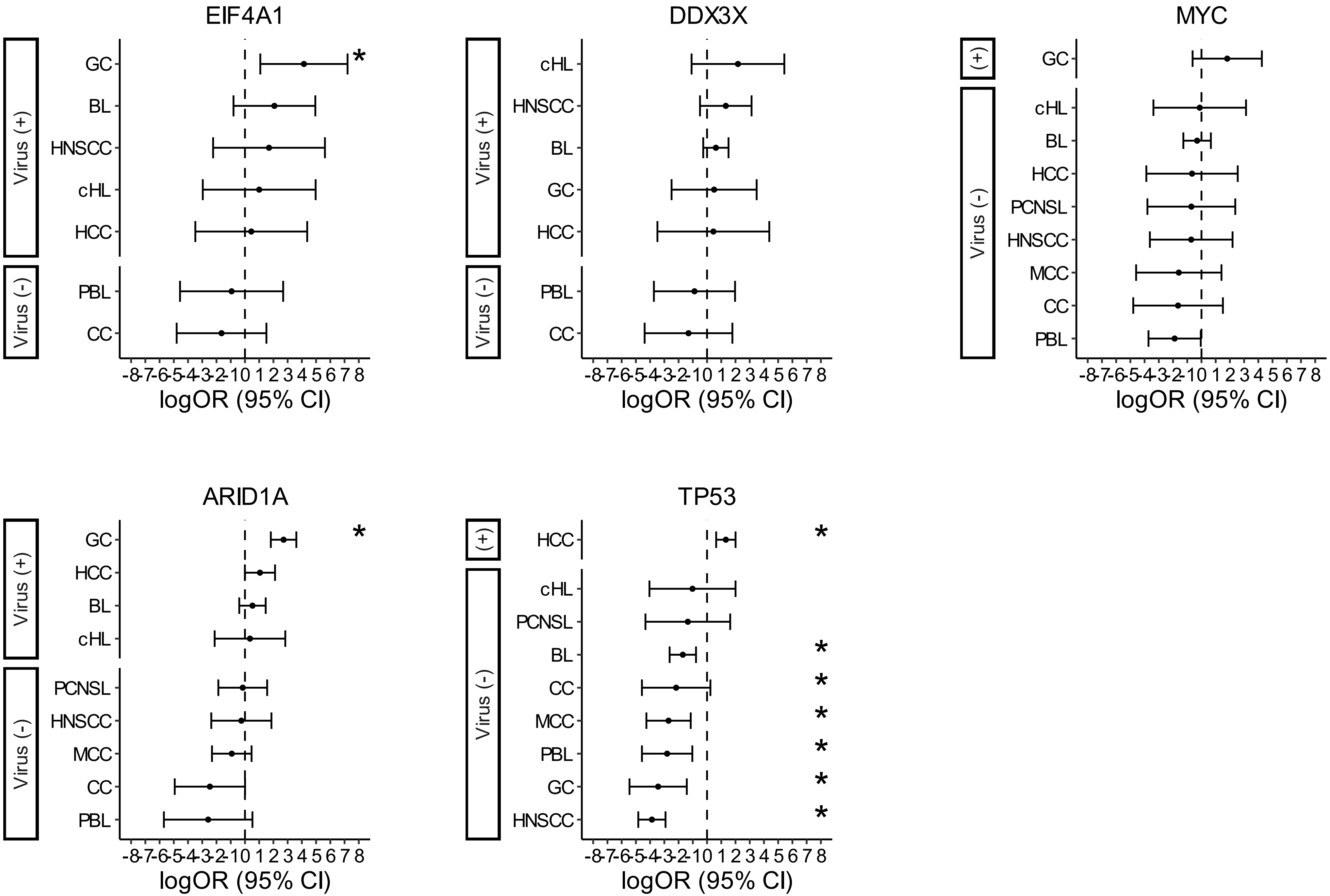
EIF4A1
Virus (+)
GC
BL
HNSCC
cHL
HCC
Virus (-)
PBL
CC
*
DDX3X
Virus (+)
cHL
HNSCC
BL
GC
HCC
Virus (-)
PBL
CC
MYC
(+)
GC
Virus (-)
cHL
BL
HCC
PCNSL
HNSCC
MCC
CC
PBL
ARID1A
Virus (+)
GC
HCC
BL
cHL
Virus (-)
PCNSL
HNSCC
MCC
CC
PBL
*
TP53
(+)
HCC
Virus (-)
cHL
PCNSL
BL
CC
MCC
PBL
GC
HNSCC
*
*
*
*
*
*
*
-8-7-6-5-4-3-2-1 0 1 2 3 4 5 6 7 8
logOR (95% CI)

S7

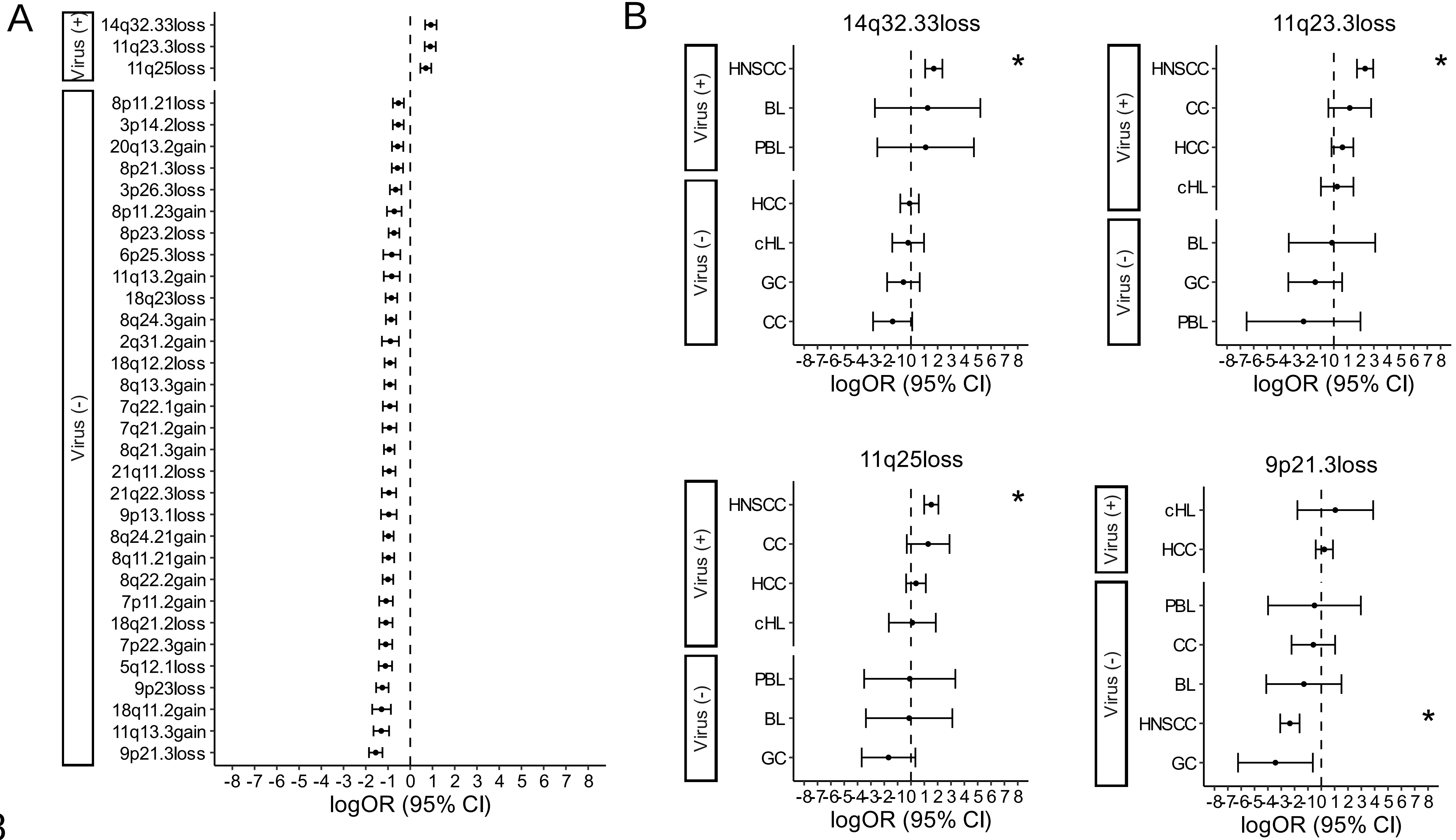
A
Virus (+)
14q32.33loss
11q23.3loss
11q25loss
Virus (-)
8p11.21loss
3p14.2loss
20q13.2gain
8p21.3loss
3p26.3loss
8p11.23gain
8p23.2loss
6p25.3loss
11q13.2gain
18q23loss
8q24.3gain
2q31.2gain
18q12.2loss
8q13.3gain
7q22.1gain
7q21.2gain
8q21.3gain
21q11.2loss
21q22.3loss
9p13.1loss
8q24.21gain
8q11.21gain
8q22.2gain
7p11.2gain
18q21.2loss
7p22.3gain
5q12.1loss
9p23loss
18q11.2gain
11q13.3gain
9p21.3loss
-8 -7 -6 -5 -4 -3 -2 -1 0 1 2 3 4 5 6 7 8
logOR (95% CI)
B
14q32.33loss
Virus (+)
HNSCC
BL
PBL
Virus (-)
HCC
cHL
GC
CC
*
logOR (95% CI)
11q23.3loss
Virus (+)
HNSCC
CC
HCC
cHL
Virus (-)
BL
GC
PBL
*
logOR (95% CI)
11q25loss
Virus (+)
HNSCC
CC
HCC
cHL
Virus (-)
PBL
BL
GC
*
logOR (95% CI)
9p21.3loss
Virus (+)
cHL
HCC
Virus (-)
PBL
CC
BL
HNSCC
GC
*
logOR (95% CI)

S8

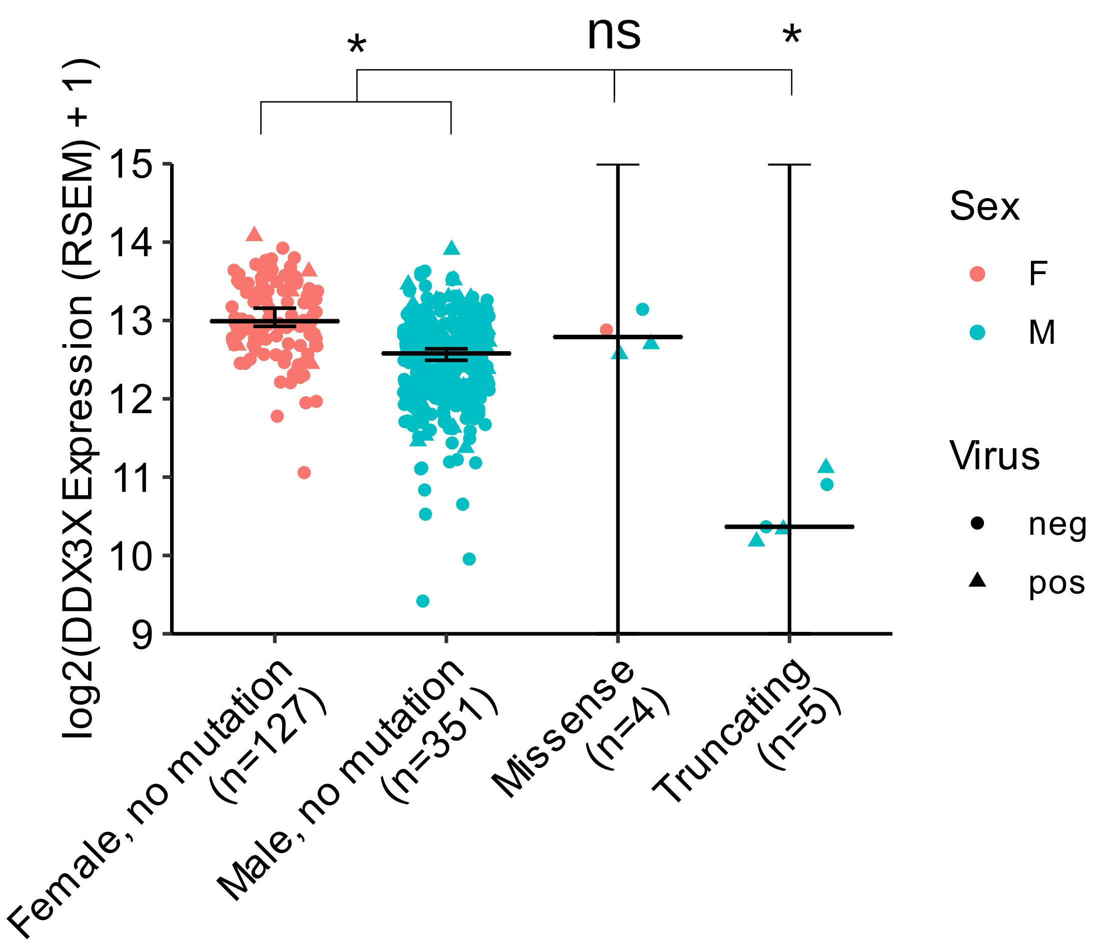

*
ns
*
log2(DDX3X Expression (RSEM) + 1)
15
14
13
12
11
10
9
Female, no mutation (n=127)
Male, no mutation (n=351)
Missense (n=4)
Truncating (n=5)
Sex
F
M
Virus
neg
pos


S9

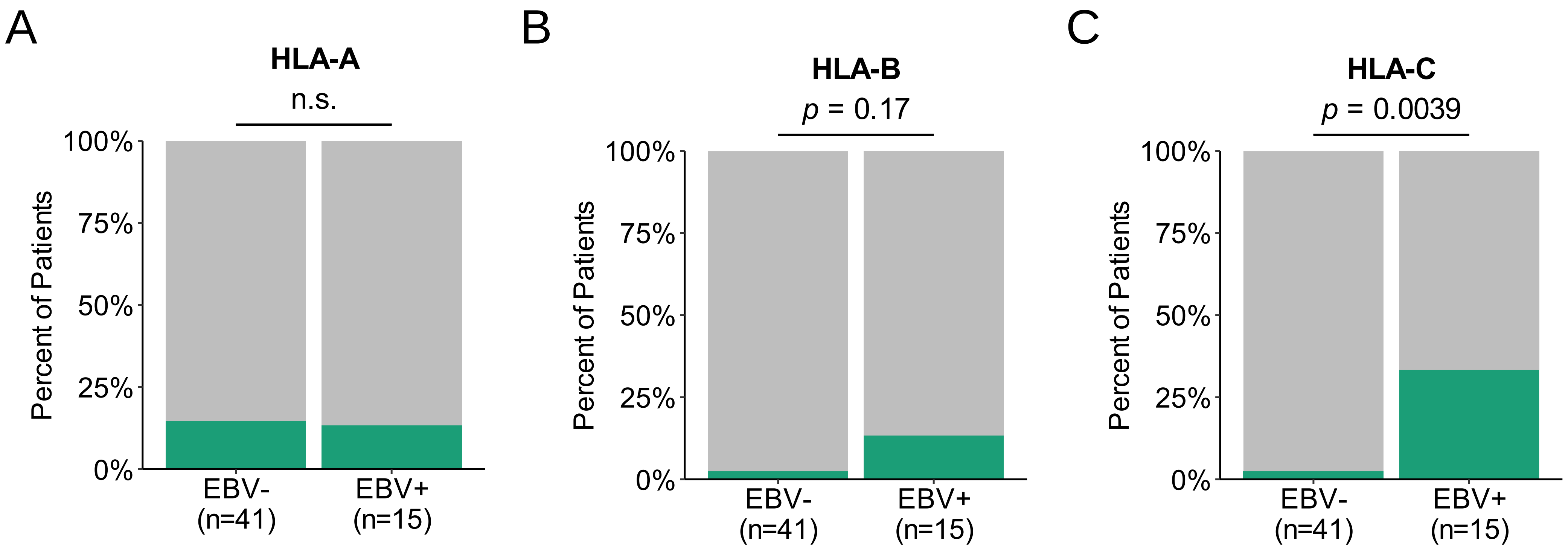

A
HLA-A
n.s.
100%
75%
50%
25%
0%
Percent of Patients
EBV-
(n=41)
EBV+
(n=15)
B
HLA-B
p = 0.17
100%
75%
50%
25%
0%
Percent of Patients
EBV-
(n=41)
EBV+
(n=15)
C
HLA-C
p = 0.0039
100%
75%
50%
25%
0%
Percent of Patients
EBV-
(n=41)
EBV+
(n=15)


S10

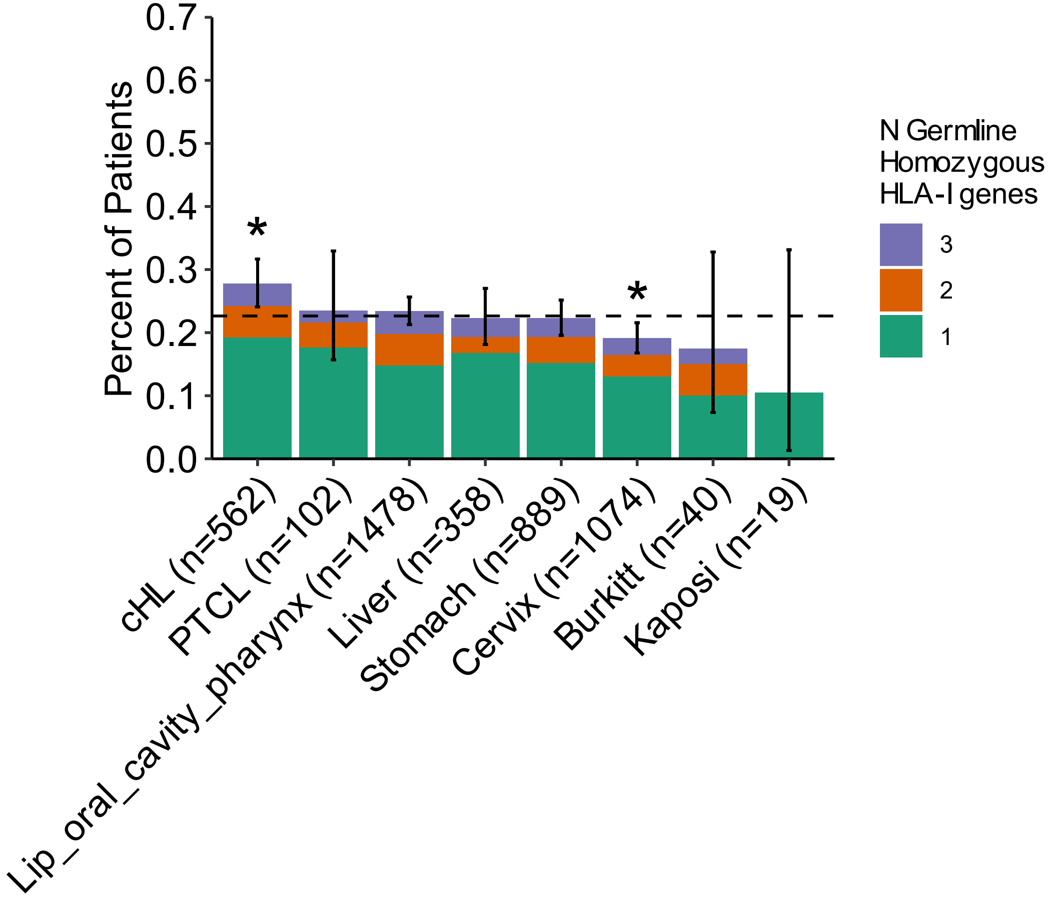

Percent of Patients
0.7
0.6
0.5
0.4
0.3
0.2
0.1
0.0
*
*
cHL (n=562)
PTCL (n=102)
Lip_oral_cavity_pharynx (n=1478)
Liver (n=358)
Stomach (n=889)
Cervix (n=1074)
Burkitt (n=40)
Kaposi (n=19)
N Germline Homozygous HLA-I genes
3
2
1


S11

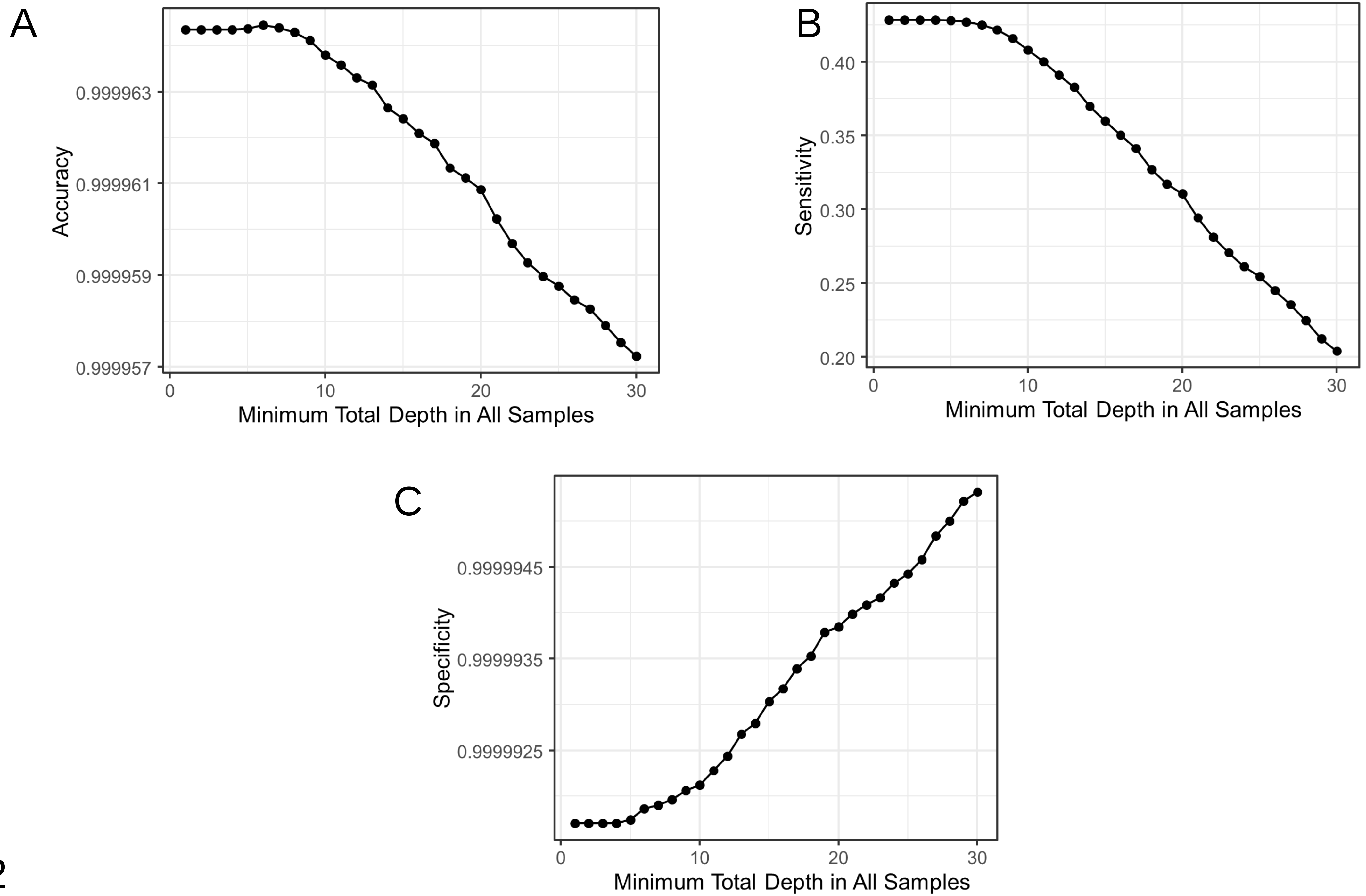
A
Accuracy
0.999963
0.999961
0.999959
0.999957
0 10 20 30
Minimum Total Depth in All Samples
B
Sensitivity
0.40
0.35
0.30
0.25
0.20
0 10 20 30
Minimum Total Depth in All Samples
C
Specificity
0.9999945
0.9999935
0.9999925
0 10 20 30
Minimum Total Depth in All Samples

S12

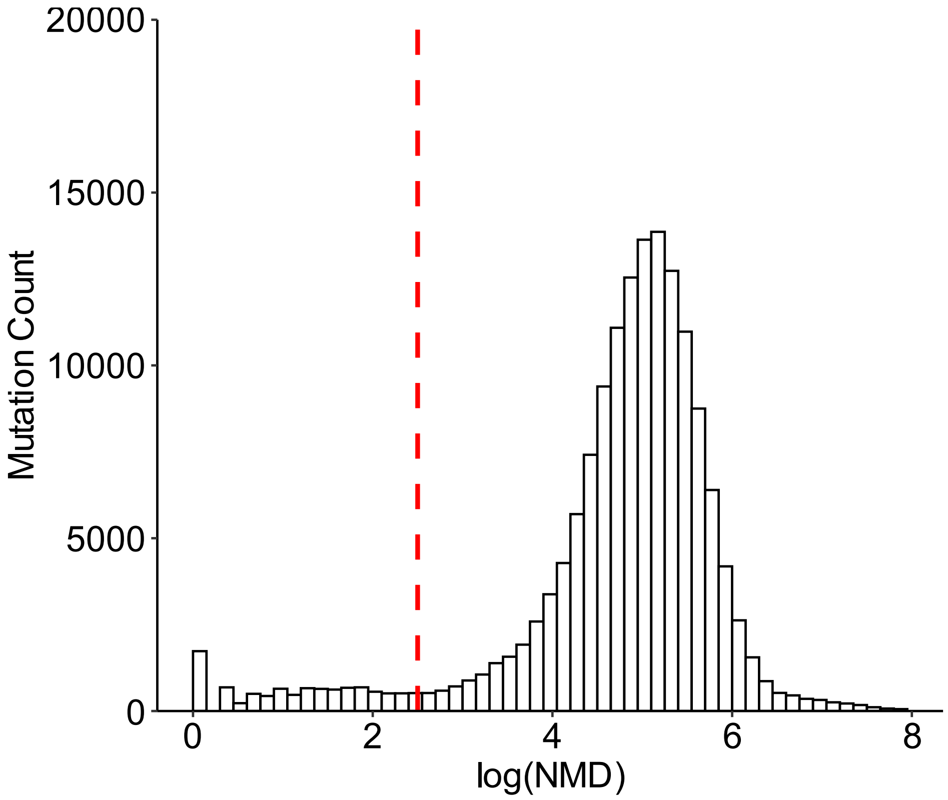


S13
